# Onset of effects of non-pharmaceutical interventions on COVID-19 worldwide

**DOI:** 10.1101/2020.09.02.20185660

**Authors:** Elisabeth L. Zeilinger, Ingo W. Nader, Dana Jomar, Clemens Zauchner

**Author notes:** these authors contributed equally (Elisabeth L. Zeilinger, Ingo W. Nader). Correspondence: Elisabeth L. Zeilinger, Faculty of Psychology, University of Vienna, Liebiggasse 5, A-1010 Vienna, Austria.

## Abstract

During the initial phase of the global COVID-19 outbreak, most countries responded with non-pharmaceutical interventions (NPIs). The effectiveness of these NPIs has been investigated with simulation studies, that rely on assumptions and by empirical studies with few countries and controversial results. However, it has not been investigated in detail how long different NPIs need to be in place to take effect, or how long they should be in place for their maximum effect to unfold. We used global data and a non-parametric machine learning model to estimate the effects of NPIs in relation to how long they have been in place. Here we show that closure and regulation of schools was the most important NPI, associated with a pronounced effect about 10 days after implementation. Restrictions of mass gatherings and restrictions and regulations of businesses were found to have a more gradual effect, and social distancing was associated with a delayed effect starting about 18 days after implementation. Generally, effects increased until about 40 to 50 days after implementation. Our results can inform political decisions regarding the choice of NPIs and how long they need to be in place to take effect.

## Background

Non-pharmaceutical interventions (NPIs) are applied by most countries around the world to reduce the risk of the COVID-19 pandemic and to slow the suspected exponential growth of infections. Exploring the impact of NPIs is crucial for gathering knowledge on effective ways to control the pandemic, and to concurrently avoid unnecessary strain on the general population, both psychologically and economically. The WHO urges that implementation of NPIs during the COVID-19 pandemic must be based on science and evidence,^1^ and the comparative analysis of the effectiveness of quarantine strategies and contexts was defined as a research gap, that should be addressed with priority.^2^ Two rapid reviews on NPIs, one on quarantine measures, one on school closures, concluded that the available evidence was very limited and lacked quality.^3,4^ Recently, there have been some great research efforts to provide comparisons of the general effectiveness of NPIs. Primarily, simulation models have been used to forecast how NPIs will most likely affect infection rates.^e.g. 5-7^ However, since multiple aspects of the COVID-19 pandemic are unclear and complex, these simulation methods have to work with parameter assumptions based on fragmented knowledge.^3^ Retrospective studies that only consider one country or region at a time in their analyses,^8,9^ suffer from the problem that multiple NPIs are introduced simultaneously. Hence, they have no means to distinguish between effects of these interventions and can only evaluate a common effect. One study compared NPIs across 11 European countries.^10^ Yet, including only few countries for analysing NPIs means low variability in the data, i.e. implemented NPIs, which can compromise and limit the results. Furthermore, existing studies using Bayesian ^e.g. 11^ or econometric methods ^e.g. 9^ assume a constant effect of NPIs over time and have no means of estimating the time NPIs need to be effective. Our study wants to address these aspects by applying a different approach. We used empirical data of 176 countries to evaluate the effectiveness of NPIs on global COVID-19 infection rates with a non-linear machine learning model. This approach allows to estimate the time it takes for each measure to show an effect on infection rates, and how long it takes to reach the maximum effect. Most countries affected by COVID-19 introduced NPIs, but they used different NPIs in different chronological order. This enables our analysis to isolate the effects of single NPIs and provide an estimation for the average effect of each NPI across all countries.

To the best of our knowledge, to date, there are no studies investigating how long it takes for an NPI to take effect, nor comparing NPIs using worldwide data. This, however, could be of great value for policy makers when considering which interventions to apply during a COVID-19 outbreak, when an effect of specific measures is to be expected, and how long it should be in place to reach maximum effect. Our results are a complementary source of information to previous studies on NPIs and may guide decision making on the careful implementation of adequate measures in relation to the COVID-19 pandemic.

## Methods

### Data sources

Three data sources were used in this study. The first dataset consisted of the time series of cumulative confirmed COVID-19 cases,^12^ collected by the Johns Hopkins University Center for Systems Science and Engineering (JHU CCSE). It is compiled from various data sources, including the WHO, and it features daily data of the total number of confirmed cases in 214 countries. The numbers are generally reported on country level, but for some large countries (Australia, Canada, and China), cases are reported by state.

The second dataset is the COVID-19 government response event dataset (CoronaNet).^13^ It is a hand-coded collection of policy announcements regarding non-pharmaceutical interventions (NPIs) of 198 countries. The data collection is performed by a large international team of researchers and includes secondary review. The policies are categorised into multiple dimensions, including 16 different types (e.g., closure and regulation of schools, restriction and regulation of businesses, social distancing), the level of government initiating the action (national, provincial, municipal), the enforcement of the policy (mandatory or voluntary), as well as special dimensions for specific types like the directionality (inbound, outbound, or both) for external border restrictions. These policies and their categorisation will be used to derive NPIs to use in the analysis (see below).

For each individual policy, the date of its implementation in the respective country is recorded, along with a free-text describing the event. The dataset lists about 31500 individual policies.

The third dataset used was the World Bank’s dataset of development indicators.^14^ In order to account for some variability between countries, we included four country-level development indicators in our analysis. In a simulation study, the difference in the effects of containment strategies was found to be most pronounced for the age group of 60+ versus the other age groups.^15^ Hence, we included the percentage of population with ages of 65 years or more,^16^ which is the indicator that comes closest in terms of age groups. Spread of COVID-19 was found to follow different patterns in urban and in rural areas,^17^ hence we included the percentage of people living in urban territories (calculated from total population^18^ and urban population^19^). Spread of airborne viruses has also been linked to atmospheric pollution like fine particulate matter (PM_2.5_) in measels,^20^ and more recently also to COVID-19.^21^ Hence, we also used the percentage of people that are exposed to PM_2.5_ air pollution values exceeding the WHO guideline value.^22^ Another factor that might influence the growth rate is the relative wealth of the inhabitants. For example, poverty has been linked to reduced compliance to shelter-in-place protocols.^23^ As a proxy, we have included the Gross Domestic Product per capita at purchasing power parity (GDP/PPP).^24^ Where available, we have downloaded the data for the last full calendar year (2019), but for countries that did not provide current data, we used the most recent data of up to 10 years prior.

### Calculating growth rates

For the analysis, the state-level information in the time series of cumulative confirmed COVID-19 cases was aggregated to country level (for Australia, Canada and China). The daily increase rate was calculated as the ratio of cumulative cases from one day to the next, for each country. To account for artefacts from variable reporting delays,^1^ we calculated a right-centred seven-day moving (geometric) mean, smoothing out weekend effects. This metric, the smoothed daily increase rate of cumulative confirmed COVID-19 cases, will be the main outcome metric for the analysis to follow. For simplicity, we will refer to this metric as the growth rate. A growth rate of one corresponds to zero new cases, i.e., no increase in cumulative cases from the previous day to the current day (within one country). A growth rate of 1.1 corresponds to a ten-percent increase.

The onset of new infectious diseases like COVID-19 is assumed to follow exponential growth,^25^_–_^27^ hence this ratio is expected to be constant over time if no NPIs are in place. Also, as the growth rate is calculated within countries, systematic over- or underreporting in a specific country (e.g., due to different testing strategies) should be effectively mitigated. We only considered days that indicated 25 or more cumulative confirmed COVID-19 cases to calculate the smoothed growth rate in order to get more stable estimates, because the confirmed cases reported in the beginning of the epidemic show considerable variation (e.g., laboratory-confirmed results may take up to one week after testing until they are reported e.g., ^28,29^). Additionally, we only used data from countries that provided at least 28 days of data. We only included countries in our analysis that the CoronaNet dataset of NPIs included data for. For a full list of countries used in the analysis, see Supplementary Table 1.

One of the assumptions of this study is that effectiveness of an NPI can be deduced from the reduction of the growth rate. This assumption is only valid in the beginning of the outbreak, as exponential growth is slowed in later phases by the implementation of NPIs and various other factors like changes in behaviour of the population in response to a global pandemic or saturation^30^. To model only the beginning, we limit our data to the first 90 days after the day that 25 cases have been reached for each country.

Because some countries changed reporting strategies in the course of the outbreak, the daily number of confirmed cases contains outliers. The most widely-known example is the day that China decided to include clinically diagnosed COVID-19 cases, alongside with laboratory tests (February 12, 2020), which led to 14108 new cases in a single day.^31^ To mitigate the effects of such outliers, the growth rate was winsorised at one for the lower bound, as well as at the 99% quantile for the upper bound: all values larger than 1.335 have been set to this value.

### Non-pharmaceutical interventions

For our analysis, we combined the individual policies listed in the CoronaNet dataset into NPIs by grouping them according to their type, the initiating country level (coded as national or sub-national), and the enforcement level (coded as mandatory, voluntary, or missing). For external border restrictions, we additionally grouped by the target direction (coded as inbound, outbound, both, or missing), as this category contained restrictions of citizens to travel abroad (outbound), as well as limitations for travellers to enter a specific country (inbound).

For each resulting NPI, the start date of the first policy in this group in each country was selected as the date of implementation (policies that did not list an implementation date were not considered). We excluded NPIs that were used in less than 20 countries, in order to get more stable and more generalisable estimates for the effects of the NPIs. This resulted in 57 NPIs, 35 of which were set on national level, and 22 on sub-national level. Regarding the enforcement level, 39 NPIs were mandatory, 18 were voluntary (1 NPI did not report any enforcement level). For a list of all NPIs used in the analysis, see Extended Data Table 1.

### Combining the datasets and calculating features

The time series of cumulative confirmed COVID-19 cases was enriched with the country-level indicators from the world bank data, providing static covariates for each country. This should enable the machine learning model to estimate country-specific variations in growth rate, if they are related to the covariates. For the minority of countries in our sample that did not provide data (6.3% for the percentage of people being 65 years or older, 1.3% for the percentage of people living in urban territories, 5.1% the percentage of people that are exposed to air pollution, and 8.9% for the GDP per capita), missing values in the world bank indicators were imputed with the median value. GDP per capita was log-transformed because of high skewness in the data.

For each NPI, a new feature was derived based on the date of first implementation (start date of the first policy in this category) and its relation to the current date in the time series of confirmed cases in the respective country. Each of those features was calculated by taking the difference of the current date in the time series and the point in time each NPI was first implemented in the respective country. This number is zero on the day an NPI was implemented, and a positive integer after the NPI came into effect, representing the number of days that an NPI has been in place. This allows to investigate the effect of an NPI depending on how long it has been in place. To also investigate the trend in the COVID-19 growth rate before the NPI was implemented, the feature was allowed to be negative, encoding the number of days before the NPI came into effect. We capped the value at −14, with all smaller (earlier) values being set to this value, hence capturing the effect also during two weeks prior to NPI implementation. For countries that did not implement a specific NPI, the feature variable was set to a value of −15, allowing the machine learning model to distinguish between countries that have put an NPI in place and those that have not. Using a feature coding like this allows a non-linear machine learning model to estimate the strength of an NPI on each individual day. Starting from 14 days prior to implementation additionally allows to assess the validity of estimated model effects (effects should only occur after implementation).

In addition, two features that capture NPI-independent, time-related changes in the growth rate have been used in the analysis. First, an absolute time scale measuring the days in relation to 11 March 2020, the day that COVID-19 has been declared a pandemic by the WHO. This should allow for capturing differences in growth rates for countries that have faced the outbreak earlier (like China where the outbreak started) or later (like African countries) during the worldwide outbreak. The second feature is a relative time scale within each country. It measures the time relative to the day that 25 cases have been reached. This should allow to capture changes in growth rates that are unrelated to NPIs but depend on the timing of the local outbreak like increasing awareness of the danger^32,33^ and related changes in behaviour that might affect the growth rate.

The final dataset for analysis included 57 NPIs, two time-related covariates that are independent from the NPIs, as well as four country-specific covariates. The dataset contains the daily growth rate for 14559 day/country-combinations from 176 countries, up to 17 August 2020.

### Fitting the machine learning model

To analyse the effect of the different NPIs, we fitted a random forest regression model.^34^ The dependent variable was the growth rate, and the features consisted of four country-specific covariates, two time-related covariates, and 57 NPIs, each representing how long a certain NPI has been in place or if it has been in place at all (as described above). Note that we do not treat this data as time series data in our analysis. Due to the exponential growth in cumulative COVID-19 cases, the growth rate is expected to be constant at a certain value if no NPIs are in place, and independent of the day before or the day after. The actual value should only depend on the set of NPIs that currently are in place in each country on a given day. Neither the actual date nor the country that the daily value of the growth rate originally stems from are used as features in the model.

The data was split into training and test set using a 60%/40% split blocked by country, i.e., the data of a specific country was either completely in the training or in the test set (and not spread across both sets). This allows the model to be validated on data from the roughly 40% of the countries that it has not been trained on. The training set was used for hyperparameter tuning. We used a random search as hyper-parameter optimization strategy,^35^ performing 100 iterations with ten-fold cross-validation within the training set. To reduce overfitting, we used a restricted hyperparameter search space. The best resulting model was refit on the complete training set, and performance was estimated on the test set.

To estimate uncertainty in the model predictions as well as in the estimated effects of the NPIs, we used bootstrapping. From the training set, we drew 100 bootstrap samples, also blocked by country, i.e., randomly choosing countries (all days of the respective country) until the size of the data set matched the original training set. We then refit the model on these bootstrap samples (without hyperparameter tuning).

Even though we fit a model to predict the COVID-19 growth rates in different countries, this is not the focus of the study. Fitting this model is merely a means to an end. We are interested in understanding how and when different NPIs affect the growth rate, and the methodology to achieve this is described in the next subsection. However, this methodology is based on the machine learning model, hence we report the fit statistics here in the Methods section. The model fit on the complete training set, as well as the bootstrap models, were able to approximate the data reasonably well, given we can only estimate an average effect over all countries. Fit statistics (Extended Data Table 2) indicate a slight overfit in the training set, but the models were able to explain about 47% of variance in the test set (on average in the bootstrap samples). Additionally, we used the models to predict the time series of growth rates for each individual country in our dataset, given the sequence of NPIs that have been chosen by that country. Extended Data Fig. 6 shows examples of reasonable fits of the models. It can clearly be seen that the models predict average effects of the NPIs in place, i.e., they have learned the average effect of the NPIs (see Results), which works well for some countries, but not so well for others (Extended Data Fig. 7). Common mistakes the models make are that they cannot predict untypically high growth rates or a resurgence of the growth rate after an initial decline (which might occur, for example, due to increased testing or because some NPIs or policies are no longer in place, e.g., if they do not have a reported end date in the CoronaNet dataset). The onset of the reduction of the growth rate is predicted slightly too early or too late in some cases, and the models cannot predict time series with untypical trends in the growth rate (Extended Data Fig. 7).

### Assessing effects of NPIs

To understand how each NPI influenced the growth rate (according to the random forest regression model), accumulated local effect (ALE) plots^36^ were used. These plots show what the model has learned about the influence of a specific feature over the whole value range of that feature. In our case, because of the way the features were designed, they show how the growth rate changes in relation to a specific NPI, from two weeks prior to implementation to 60 days after. They show the main effect of a feature at a certain value, compared to the average prediction, and they are centred at zero. Compared to similar plots like partial dependence plots, ALE plots do not suffer from the extrapolation problem, are not biased by the omitted variable phenomenon, and are well-suited to handle correlated features.^37^ ALE plots do not provide a means to assess uncertainty, but they are much faster to calculate compared to their alternatives. This allows to use bootstrap sampling to assess uncertainty. The plots show what the model has learned and do not depend on the dataset that is used to draw the plots (except for random fluctuation in the feature range). Hence, we have bootstrapped the model training process (as described above) and we show ALE plots of the bootstrapped models to allow assessment of uncertainty. The plots are constructed using the full dataset and show the individual bootstrap results, the median effect over all bootstraps, as well as the model which has been estimated on the complete training set.

All analyses were carried out using R,^38^ mainly using the packages mlr3,^39^ ranger,^34^ iml,^40^ wbstats,^14^ and ggplot2.^41^ All calculations were performed on an IBM POWER8 platform with 80 threads on 14 physical cores. The code for this analysis was subject to code review among the authors.

### Data availability

All data used in this analysis is publicly available. The time series of cumulative confirmed COVID-19 cases was downloaded from https://data.humdata.org/dataset/novel-coronavirus2019-ncov-cases, the government response dataset (CoronaNet) was downloaded from https://github.com/saudiwin/corona_tscs, and the Word Bank’s development indicators were downloaded via the R package wbstats.^14^ The datasets generated during and/or analysed during the current study are available from the corresponding author on reasonable request.

## Results

The most important NPIs to reduce the growth rate during the COVID-19 outbreak in 2019/2020, as identified by the machine learning model in the context of our data, were closure and regulation of schools, restrictions of mass gatherings, social distancing, and restrictions and regulation of businesses, all with mandatory enforcement on national level (Fig. 1). The first effects started to show about 10 days after implementation, and effects generally lasted until about 40 to 50 days after implementation.

**Fig. 1.**
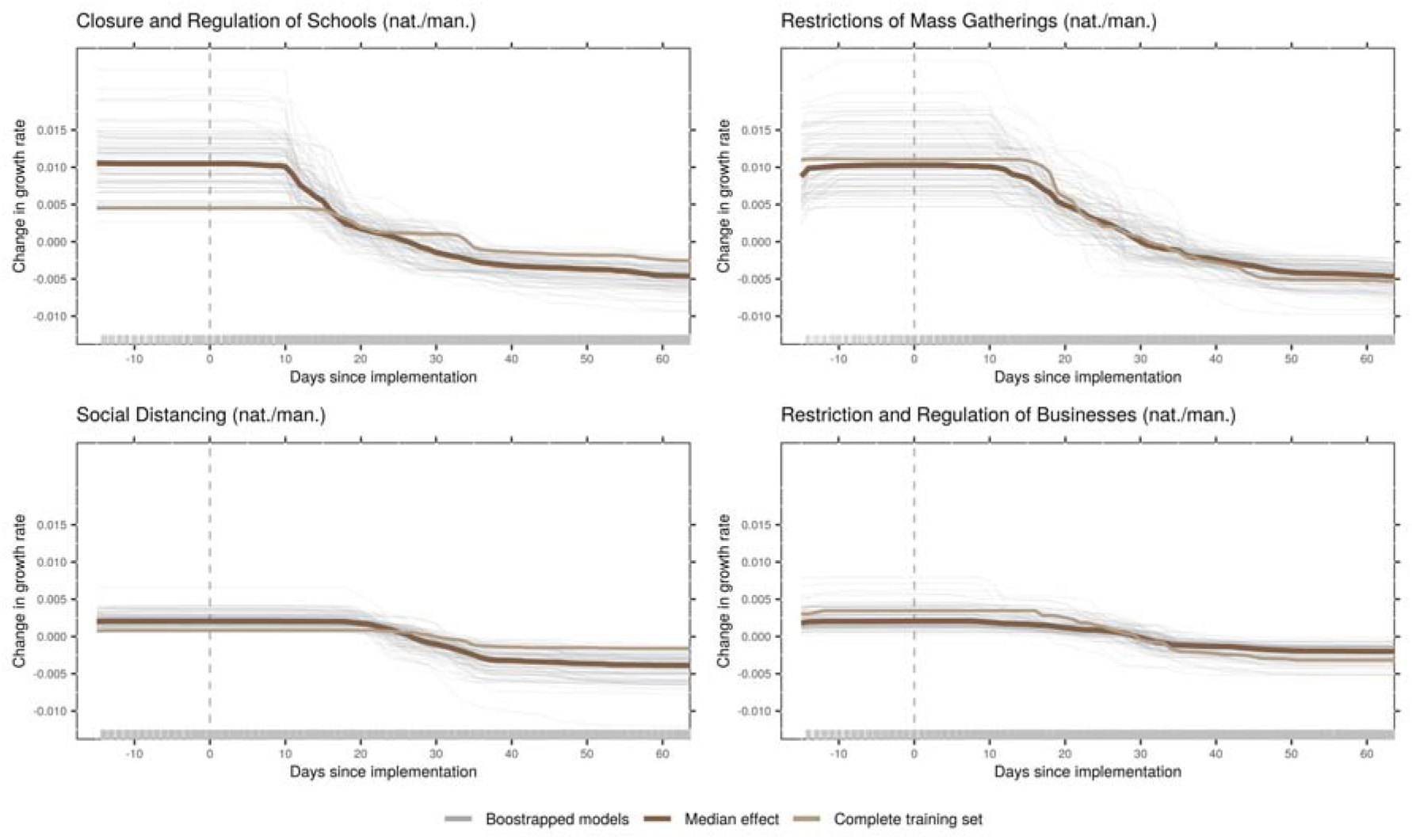
Effects of NPIs as identified by the model. The panels of this figure show predicted changes in the growth rate given how long a certain NPI has been in place. These are the main effects (ALE plots) for the most important NPIs, as identified by the model. The dark brown line is the median effect over all bootstrap samples, grey lines depict individual bootstrap samples, and the light brown line represents the complete training set. The day of implementation is marked with a vertical, dashed line. Plots show a time frame of two weeks prior to the measure to 60 days after implementation.

Closure and regulation of schools was associated with a rather distinct drop in the predicted growth rate at around 10 days after implementation of the NPI (Fig. 1). On average over the different countries in the sample, this NPI was introduced relatively timely within each country (Extended Data Fig. 1).

Restrictions of Mass Gatherings were associated with a more gradual decrease starting around 10 days after implementation. This NPI was introduced rather early within each country (Extended Data Fig. 1). Relatedly, the growth rate was still relatively high, compared to other NPIs (Extended Data Table 1).

Social Distancing includes measures like mask wearing, keeping a minimum distance to other individuals, and banning visits to hospitals or other institutions. This NPI was introduced relatively late within each country (Extended Data Fig. 1). The reduction of the growth rate started around 18 days after implementation (Fig. 1), with some bootstrap results showing first effects at around 10 days. The maximum decrease was reached 40 days after implementation.

Restriction and Regulation of Businesses started to show slight effects around 10 days after implementation. The total decrease was rather small, compared to other NPIs and to the random variation in the bootstrap samples (Fig. 1).

Further NPIs used in the analysis were associated with only minor effects (see Extended Data Fig. 2-5 for plots of all NPIs and Supplementary Information for further results and discussion).

Time-related, NPI-independent effects were larger compared to effects associated with NPIs. The average growth rate exhibits a sharp drop around 20 days after the WHO declared COVID-19 a pandemic (Fig. 2). Within countries, the model identified a decrease in growth rate starting from around two weeks after 25 cases have been reached, unrelated to NPIs. The relations of country-specific covariates and predicted growth rate were small and hardly exceeded the random variation in the bootstrap samples (Figure 3; see also Supplementary Information).

**Fig. 2.**
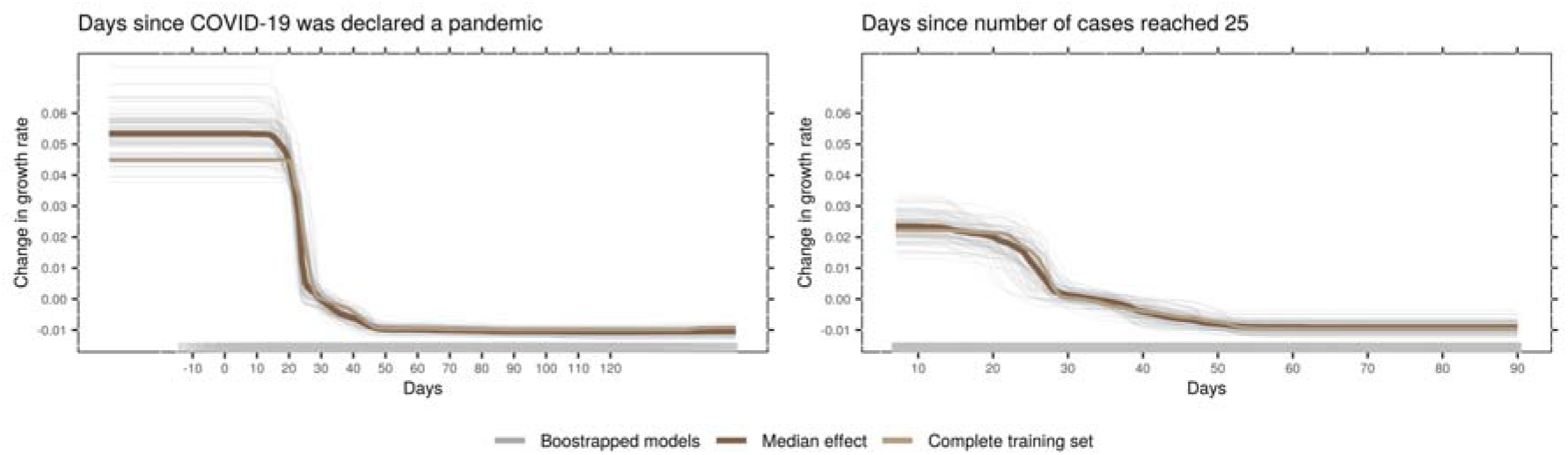
Time-related, NPI-independent effects as identified by the model. The panels show the predicted growth rate given the two time-related, but NPI-independent covariates that were used in the analysis. The panel on the left shows an absolute time scale, indicating the timing in the global COVID-19 outbreak (measured in relation to March 11, 2020, the day the WHO declared COVID-19 a pandemic). The panel on the right is a relative time scale within each country. It indicates how recent an outbreak is within that country (measured in relation to the day that 25 cumulative COVID-19 cases were reached). The dark brown line is the median effect over all bootstrap samples, grey lines depict individual bootstrap samples, and the light brown line represents the complete training set.

**Fig. 3.**
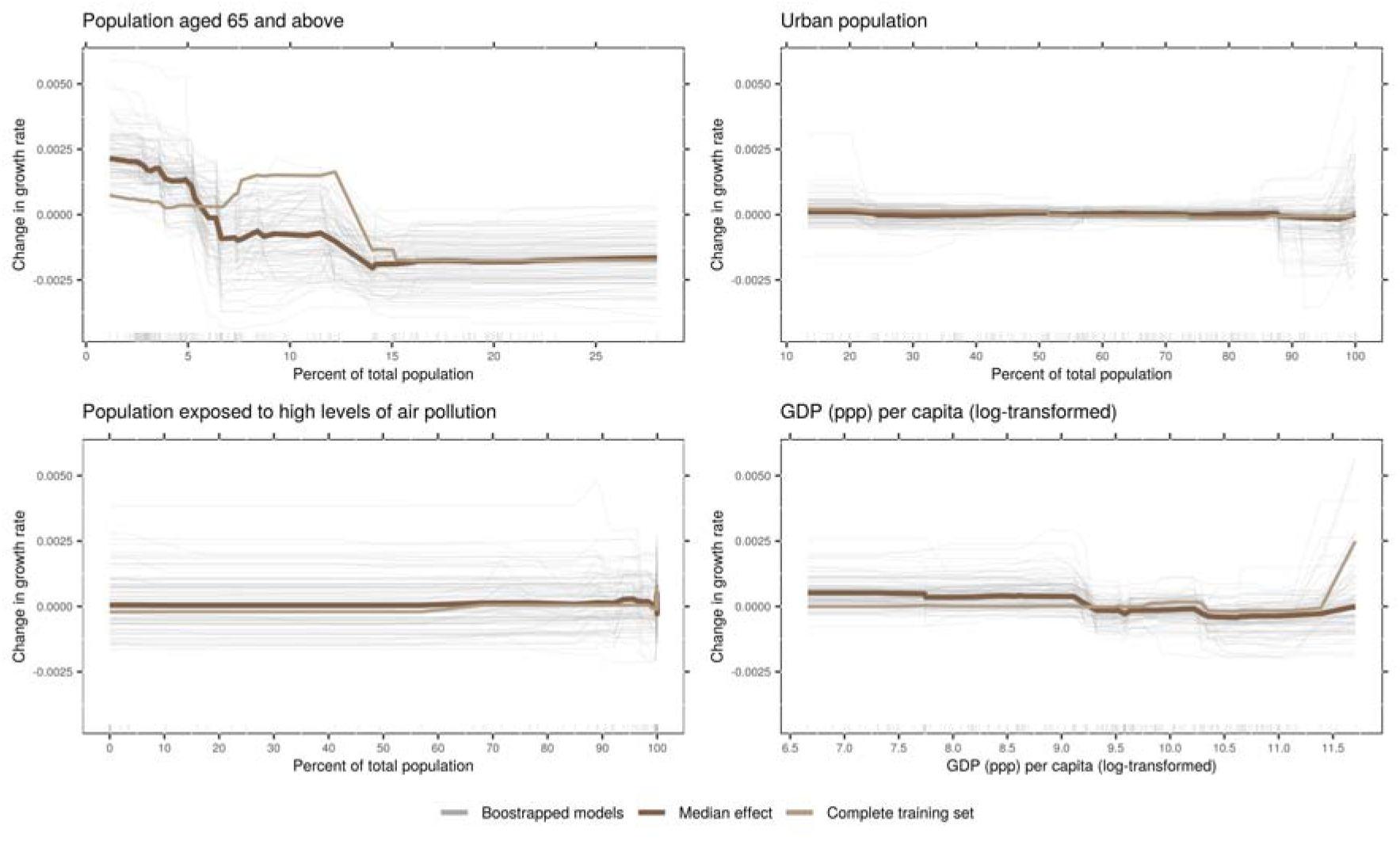
Country-specific covariates and their effects as identified by the model. The panels show the predicted growth rate in relation the four country-specific covariates included in the model. The GDP at purchasing parity power (ppp) per capita was log-transformed to account for the high skewness. The dark brown line is the median effect over all bootstrap samples, grey lines depict individual bootstrap samples, and the light brown line represents the complete training set.

## Discussion

This study investigated the effects of NPIs on the COVID-19 growth rate during the initial phase of the outbreak in 2019/2020. To complement other approaches like simulation studies, we used empirical data to train a non-linear machine learning model and examined the relationship of each NPI with the model’s prediction for the growth rate. We identified four NPIs, all with mandatory enforcement, as most effective: closure and regulation of schools, restrictions of mass gatherings, social distancing, and restrictions and regulation of businesses.

Our approach advances to the current research by investigating how the growth rate changes in the first days and weeks after NPI implementation, providing an estimate of how long it takes for an NPI to take effect. This timing varied between the four most important NPIs, from around 10 to 18 days. Another study that attempted to estimate the onset of effects, which was limited to weekly granularity and a Chinese subpopulation, found similar timing with no effects in the first week, and steadily increasing effects from weeks two to five for the NPIs they considered (emergency declaration, travel ban, and home isolation).^9^ We found that effects of NPIs generally lasted about until 40 to 50 days after implementation. After this time, the maximum possible reduction of the growth rate associated with an NPI has been reached. However, our analysis cannot give an estimate about the possible rise of the growth rate once NPIs are lifted.

Closure and regulation of schools was associated with the most pronounced and earliest drop in the growth rate. The large effect we identified contradicts a rapid review, concluding that school closures might not be effective.^4^ However, this conclusion was based on only one simulation study relying on schools data from the UK and transmission dynamics data from China.^42^ Our results are supported by a recent US study^43^ that found noticeable effects of school closures, although the methodology did not allow for isolating the effect of school closures from the effect of other concurrently present NPIs.

For the other NPIs discussed above, a more gradual drop was observed. A potential reason for this could be a change in intensity of the NPIs during the COVID-19 outbreak. In the beginning, only large gatherings were cancelled, only a few businesses were closed, and social distancing meant maintaining some distance. As the outbreak’s severity increased, so did the NPIs, with more businesses closed, more and smaller gatherings cancelled, and wearing mask becoming increasingly common.

Social distancing showed late effects compared to other NPIs. One possible explanation is that, despite mandatory enforcement, the effectiveness depends on people’s willingness to adhere to this NPI. In face of the perceived negative consequences,^44^ people might have been reluctant in the beginning, but more willing to follow these rules as the outbreak became increasingly more severe and threatening.^32^

In addition to the effects of the NPIs, the model identified strong time-related, but NPI-independent effects, related to the global spread of COVID-19, as well as to the increasing severity within the respective country. An explanation of the global time effect might be that countries affected later could have been better prepared to react to the new disease. The within-country effect might be related to behaviour changes (e.g. hand hygiene) due to the perceived severity of the threat posed by COVID-19,^32^ as the number of cases grows and news coverage is high.

When estimating average effects of NPIs on a global scale, it must be acknowledged that different countries are affected differently by COVID-19.^45^ It has been found to follow different patterns in urban and rural areas,^17^ and spread has been linked to atmospheric pollution.^21^ The effects of NPIs have been found to differ by age^15^ and relative wealth or poverty.^23^ In our analysis, we included country-level covariates as proxies for these differences, but the effects associated with these proxies were only minor. In our opinion, this does not contradict the findings above, but rather shows that between-country differences cannot be modelled effectively by country-level indicators. More fine-grained, individual level data would be necessary, as even something as personal as risk perception^33^ and fear^32^ can influence the adoption of preventative health behaviours and therefore the effectiveness of NPIs.

A few limitations have to be called out for our study. First, it is an observational study, not an experimental design. Confounding factors, e.g. environmental parameters like climate, which generally influence viral transmission^46^ and which vary between countries, cannot be ruled out to influence results. Moreover, the analysis is unable to distinguish between correlation and causation, which makes interpretation of effects difficult. These are limitations that are shared by all observational studies.^e.g. 8^ Another concern is data quality. Confirmed cases are reported daily, but with varying reporting delays.^1,28^ Some countries have changed the case definition for COVID-19 during the outbreak (e.g. China^47^), leading to artificial spikes in the time series of cumulative cases. We tried to mitigate these effects by using a moving average over a full week and by reducing the effect of outliers through winsorisation. Different testing strategies between countries, or systematic underreporting of cases might pose another problem, which we tried to combat by using relative changes within countries (growth rate) as our outcome metric. Furthermore, despite the efforts of the CoronaNet project to standardise NPIs across countries, the reported NPIs are not mutually exclusive, and the individual policies of the countries that are summarised under a certain NPI are sometimes diverse, which might impair the model’s ability to estimate an effect. Even in the face of these data-quality-related problems, the model still explained almost half of variance in the data.

Despite all limitations mentioned above, we strongly believe that for examining the impact of NPIs on the COVID-19 threat, it is vital to apply diverse methods and different perspectives to gain new insights. In addition to existing empirical studies, our machine-learning-based approach permits to non-parametrically estimate how long it takes for an NPI to affect the growth rate, which has not been studied in detail, yet. We believe that our approach adds knowledge from a different point of view about COVID-19, which may facilitate the evidence-based use of NPIs, as the WHO calls for.^1^

## Data Availability

All data used in this analysis is publicly available. The time series of cumulative confirmed COVID-19 cases was downloaded from https://data.humdata.org/dataset/novel-coronavirus-2019-ncov-cases, the government response dataset (CoronaNet) was downloaded from https://github.com/saudiwin/corona_tscs, and the Word Banks development indicators were downloaded via the R package wbstats.31 The datasets generated during and/or analysed during the current study are available from the corresponding author on reasonable request.

## Acknowledgements

The authors would like to thank the IT Power Services GmbH, especially Klaus Haderer, Werner Höger, and Michael Petroni, for their commitment to contribute to the scientific process and for making this study possible.

## Author’s Contributions

ELZ and IWN conceived the study. ELZ contributed to drafting and finalising the manuscript, performed the literature search, and supervised the analysis. IWN was responsible for data preparation and data analysis, and contributed to drafting and finalising the manuscript. DJ contributed to data preparation and data analysis, performed code review and did proof reading. CZ contributed to data analysis, performed code review and did proof-reading. All authors contributed to the interpretation of data and results, reviewed and revised the manuscript and approved the manuscript in its final version.

## Extended Data

**Extended Data Table 1.**
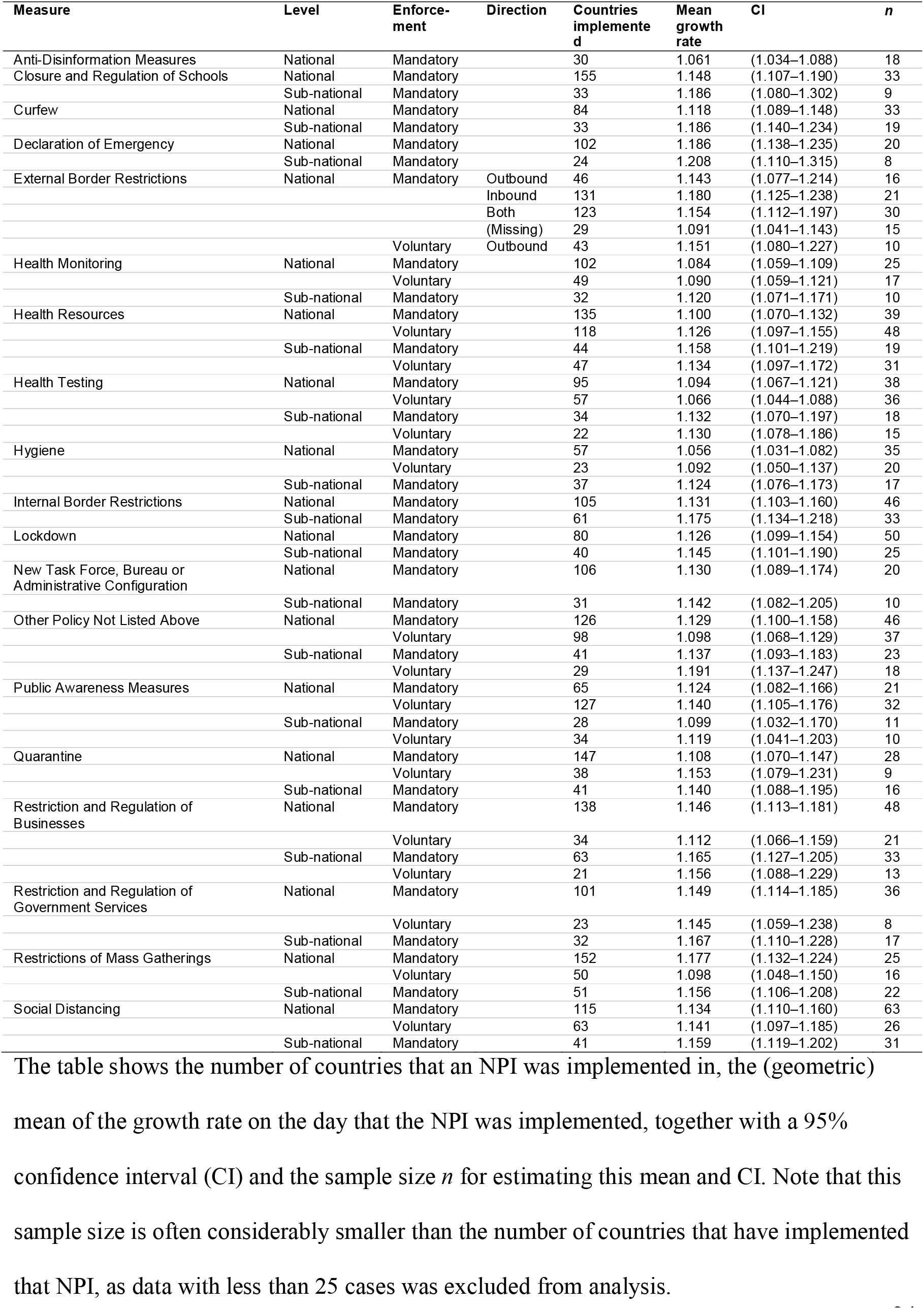
NPIs used in the analysis and growth rate at implementation.

**Extended Data Table 2.**
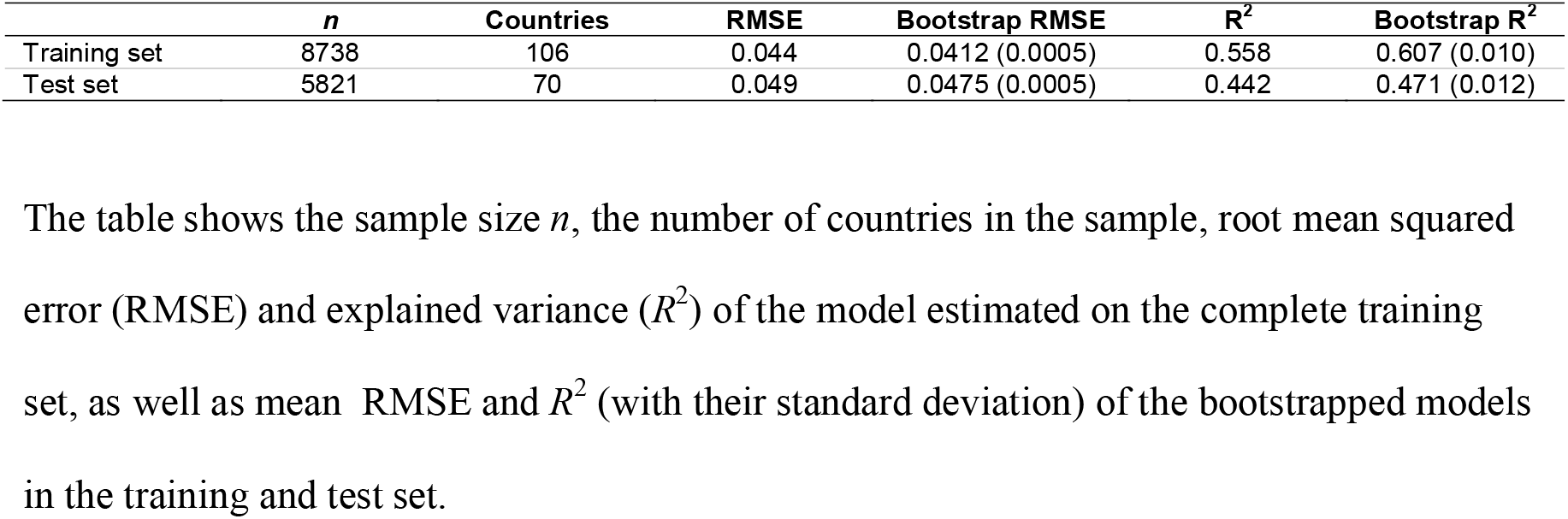
Fit statistics.

|  | **$n$** | **Countries** | **RMSE** | **Bootstrap RMSE** | **$R^2$** | **Bootstrap $R^2$** |
| --- | --- | --- | --- | --- | --- | --- |
| Training set | 8738 | 106 | 0.044 | 0.0412 (0.0005) | 0.558 | 0.607 (0.010) |
| Test set | 5821 | 70 | 0.049 | 0.0475 (0.0005) | 0.442 | 0.471 (0.012) |

The table shows the sample size $n$ , the number of countries in the sample, root mean squared error (RMSE) and explained variance ( $R^2$ ) of the model estimated on the complete training set, as well as mean RMSE and $R^2$ (with their standard deviation) of the bootstrapped models in the training and test set.

**Extended Data Fig. 1.**
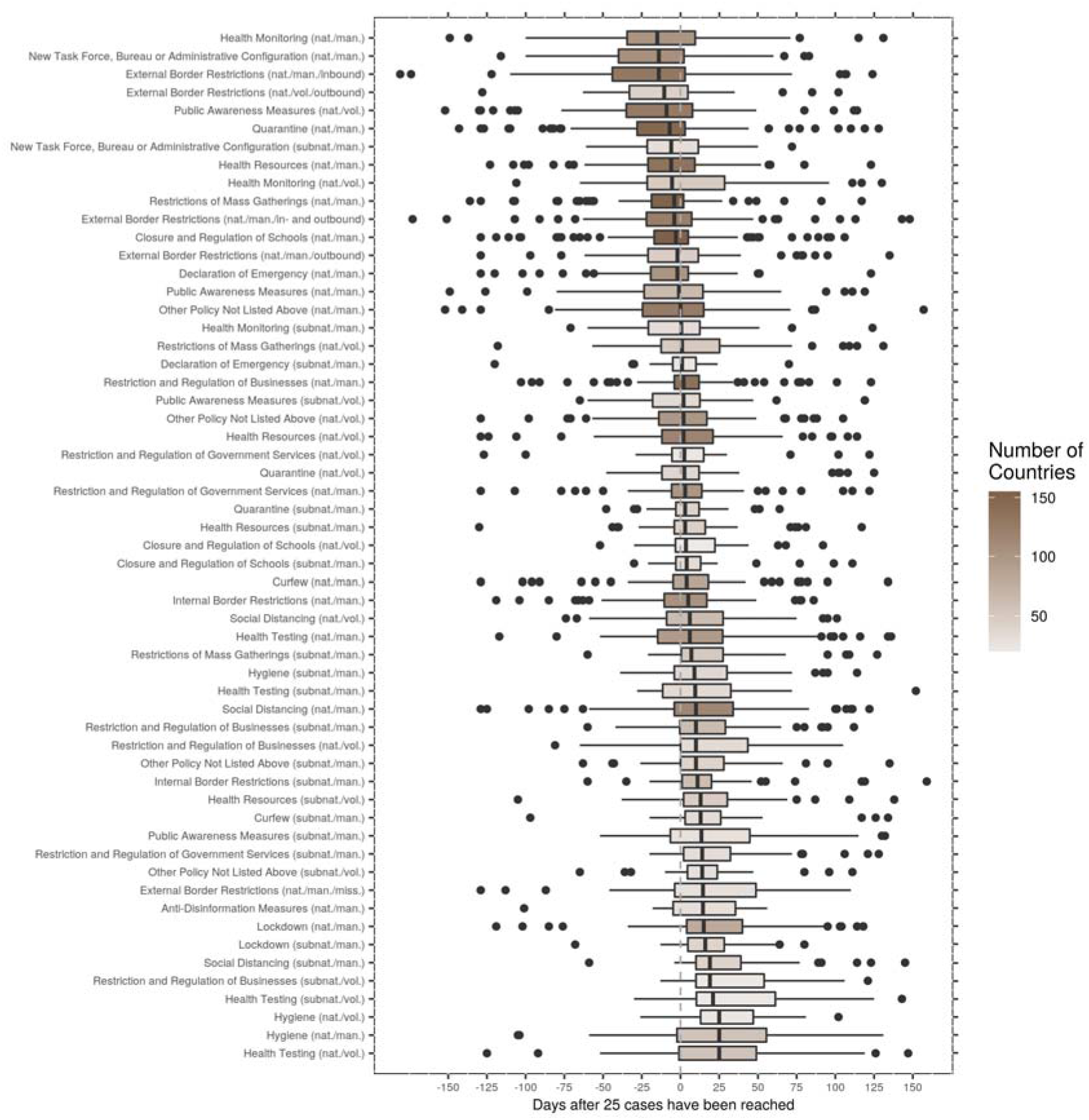
Time point of NPI implementation. The figure shows the time point at which a certain NPI was implemented, in relation to the day when a cumulative number of 25 confirmed COVID-19 cases was reached. Each data point represents one country, and the boxplots show the distribution of when these countries implemented the NPI. Colour coding corresponds to the number of countries that have implemented a specific NPI.

**Extended Data Fig. 2.**
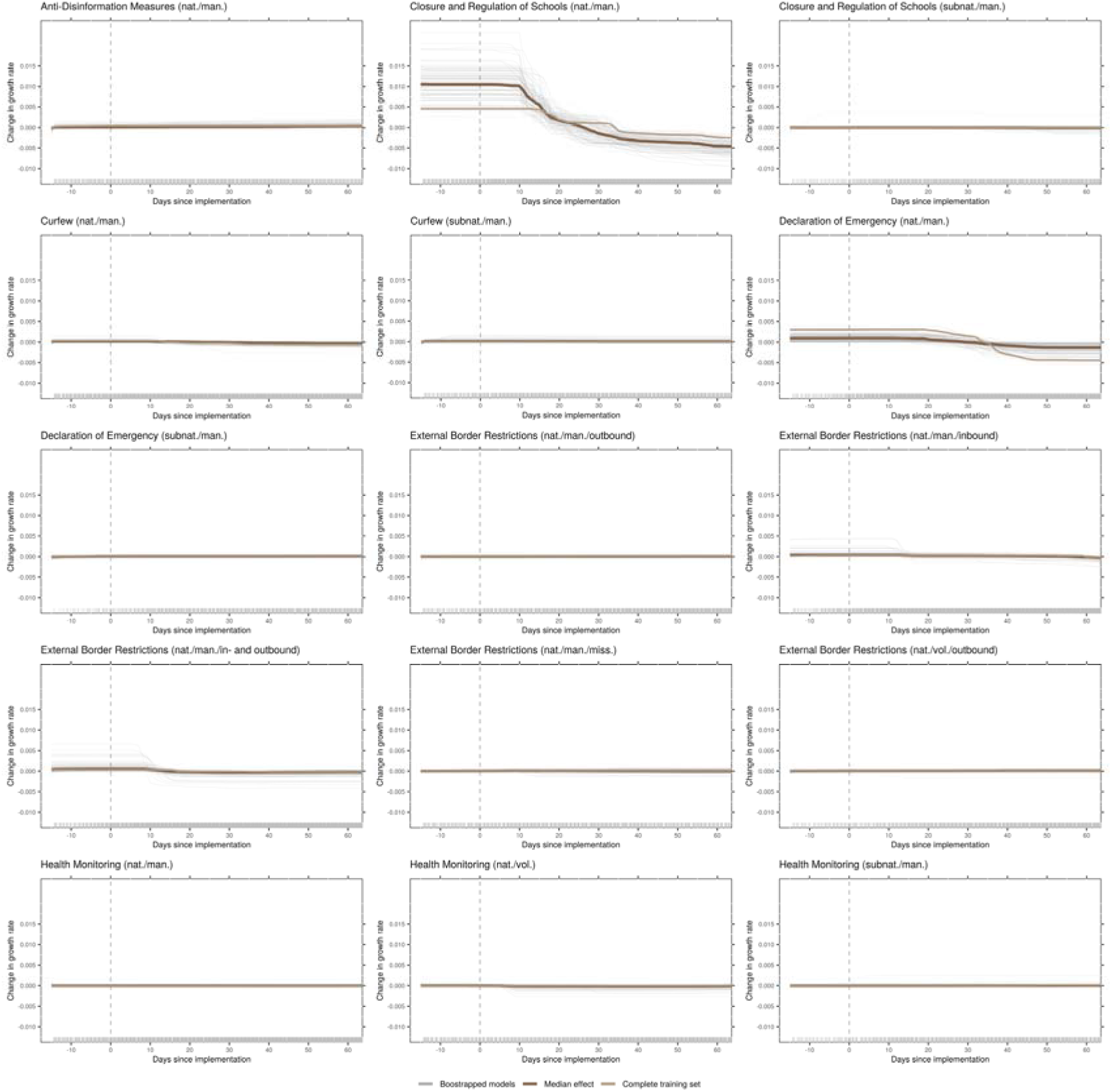
Effects of all NPIs as identified by the model (part 1) The panels of this figure show predicted changes in the growth rate given how long a certain NPI has been in place. These are the main effects (ALE plots) for the most important NPIs, as identified by the model. The dark brown line is the median effect over all bootstrap samples, grey lines depict individual bootstrap samples, and the light brown line represents the complete training set. The day of implementation is marked with a vertical, dashed line. Plots show a time frame of two weeks prior to the measure to 60 days after implementation.

**Extended Data Fig. 3.**
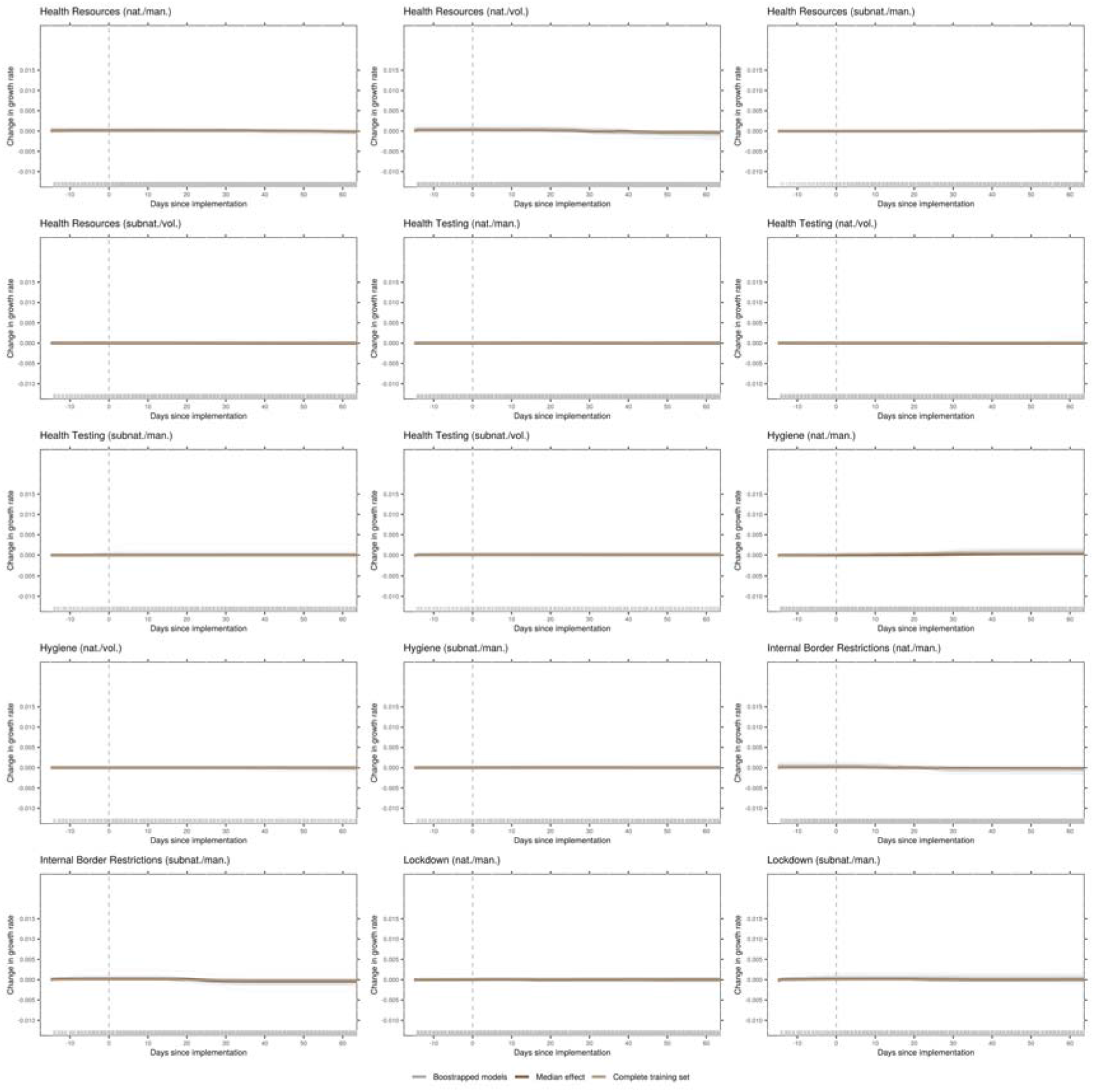
Effects of all NPIs as identified by the model (part 2) The panels of this figure show predicted changes in the growth rate given how long a certain NPI has been in place. These are the main effects (ALE plots) for the most important NPIs, as identified by the model. The dark brown line is the median effect over all bootstrap samples, grey lines depict individual bootstrap samples, and the light brown line represents the complete training set. The day of implementation is marked with a vertical, dashed line. Plots show a time frame of two weeks prior to the measure to 60 days after implementation.

**Extended Data Fig. 4.**
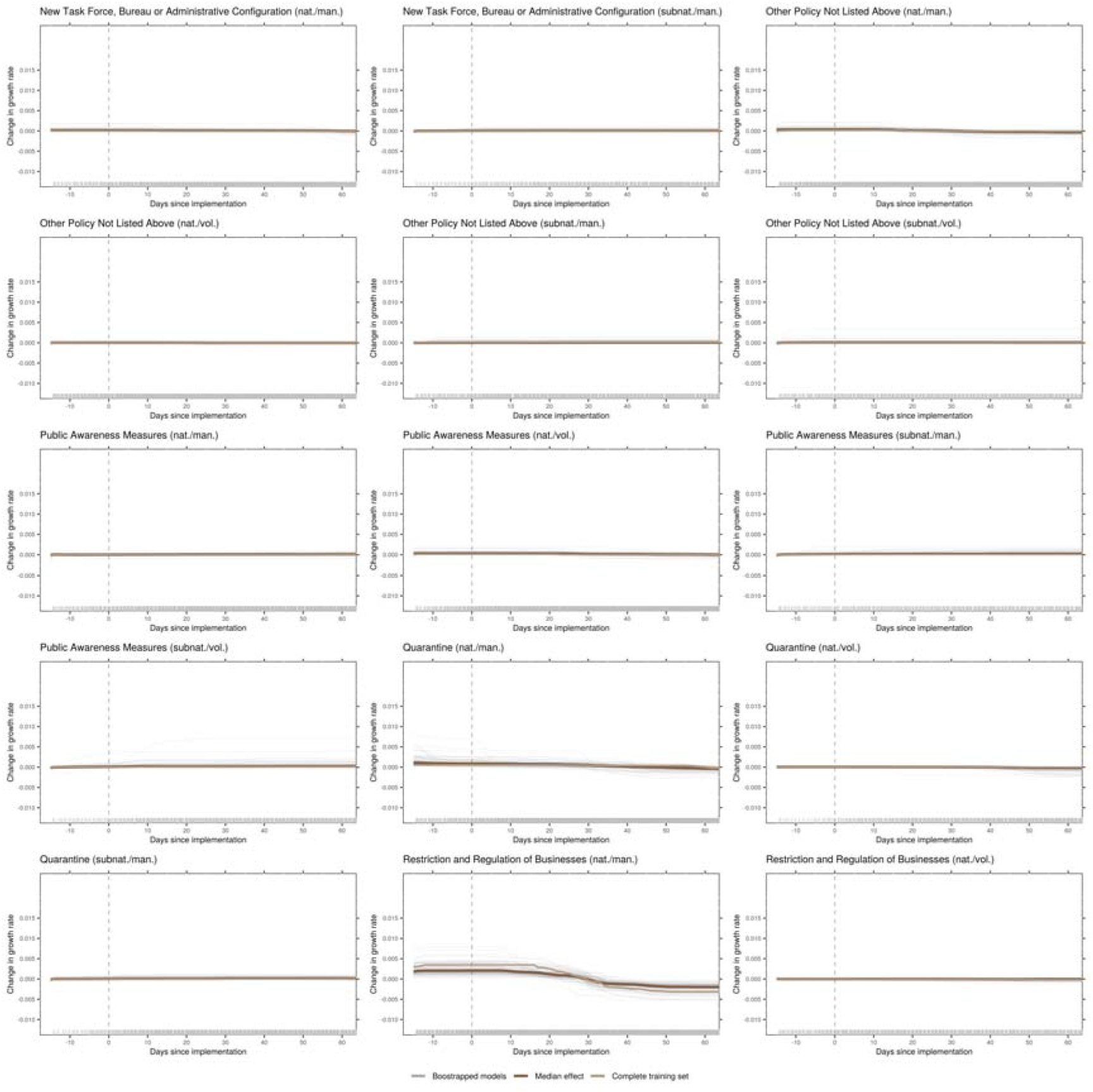
Effects of all NPIs as identified by the model (part 3) The panels of this figure show predicted changes in the growth rate given how long a certain NPI has been in place. These are the main effects (ALE plots) for the most important NPIs, as identified by the model. The dark brown line is the median effect over all bootstrap samples, grey lines depict individual bootstrap samples, and the light brown line represents the complete training set. The day of implementation is marked with a vertical, dashed line. Plots show a time frame of two weeks prior to the measure to 60 days after implementation.

**Extended Data Fig. 5.**
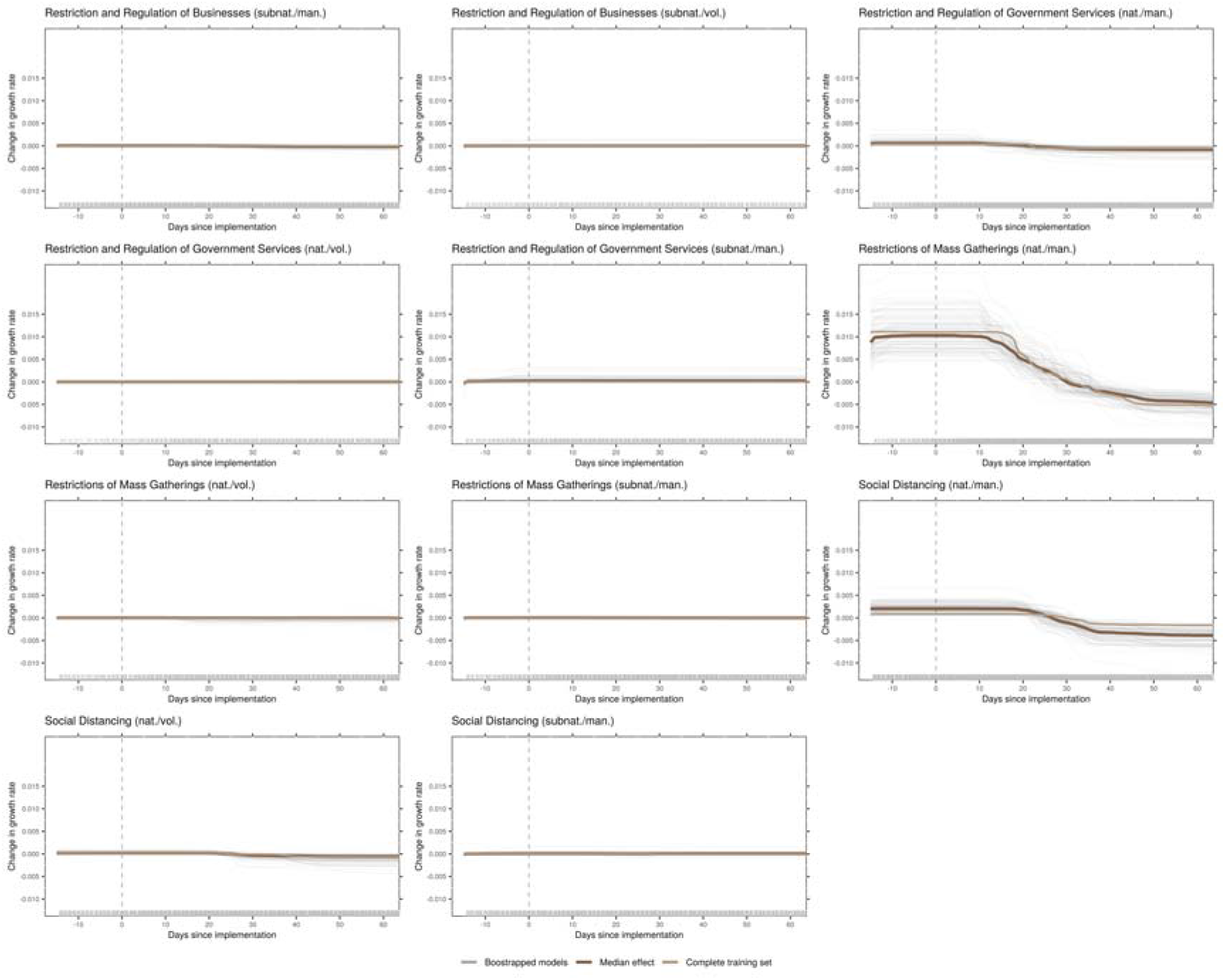
Effects of all NPIs as identified by the model (part 4) The panels of this figure show predicted changes in the growth rate given how long a certain NPI has been in place. These are the main effects (ALE plots) for the most important NPIs, as identified by the model. The dark brown line is the median effect over all bootstrap samples, grey lines depict individual bootstrap samples, and the light brown line represents the complete training set. The day of implementation is marked with a vertical, dashed line. Plots show a time frame of two weeks prior to the measure to 60 days after implementation.

**Extended Data Fig. 6.**
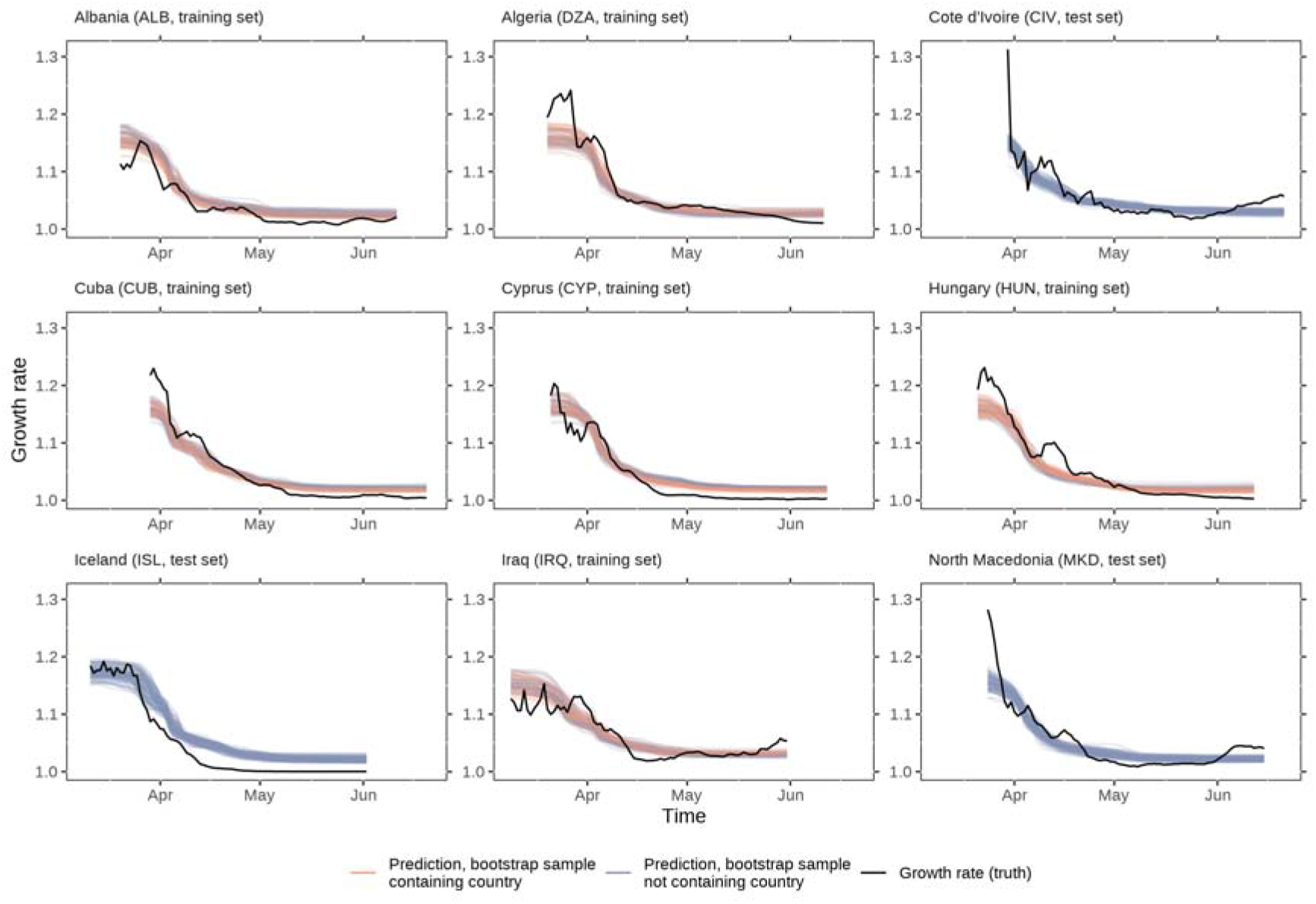
Country-specific growth rates and well-approximating predictions. The figure shows some examples of countries that the models’ predictions were reasonably accurate. The models were only able to learn average effects of the NPIs over all countries, hence they were not able to predict untypically high (e.g., DZA, CIV, MKD) or low (e.g., ALB) growth rates in the beginning of the outbreak. They were also not capable to predict rising growth rates, neither in the mid-sections of the time series (e.g., HUN, MKD) nor towards the end (e.g., CIV, IRQ, MKD). Another observation that can be made is that the models were not always able to learn the full extent in the reduction of the growth rate, as the predicted growth rates are higher than the actual growth rates towards the end of the time series (e.g., DZA, CYP, ISL). Countries that were part of the training set contain bootstrap estimates of the predictions where the respective country was part of the bootstrap sample or not (indicated by the colour of the bootstrap estimates), countries that were in the test set contain only bootstrap estimates that did not contain the respective country.

**Extended Data Fig. 7.**
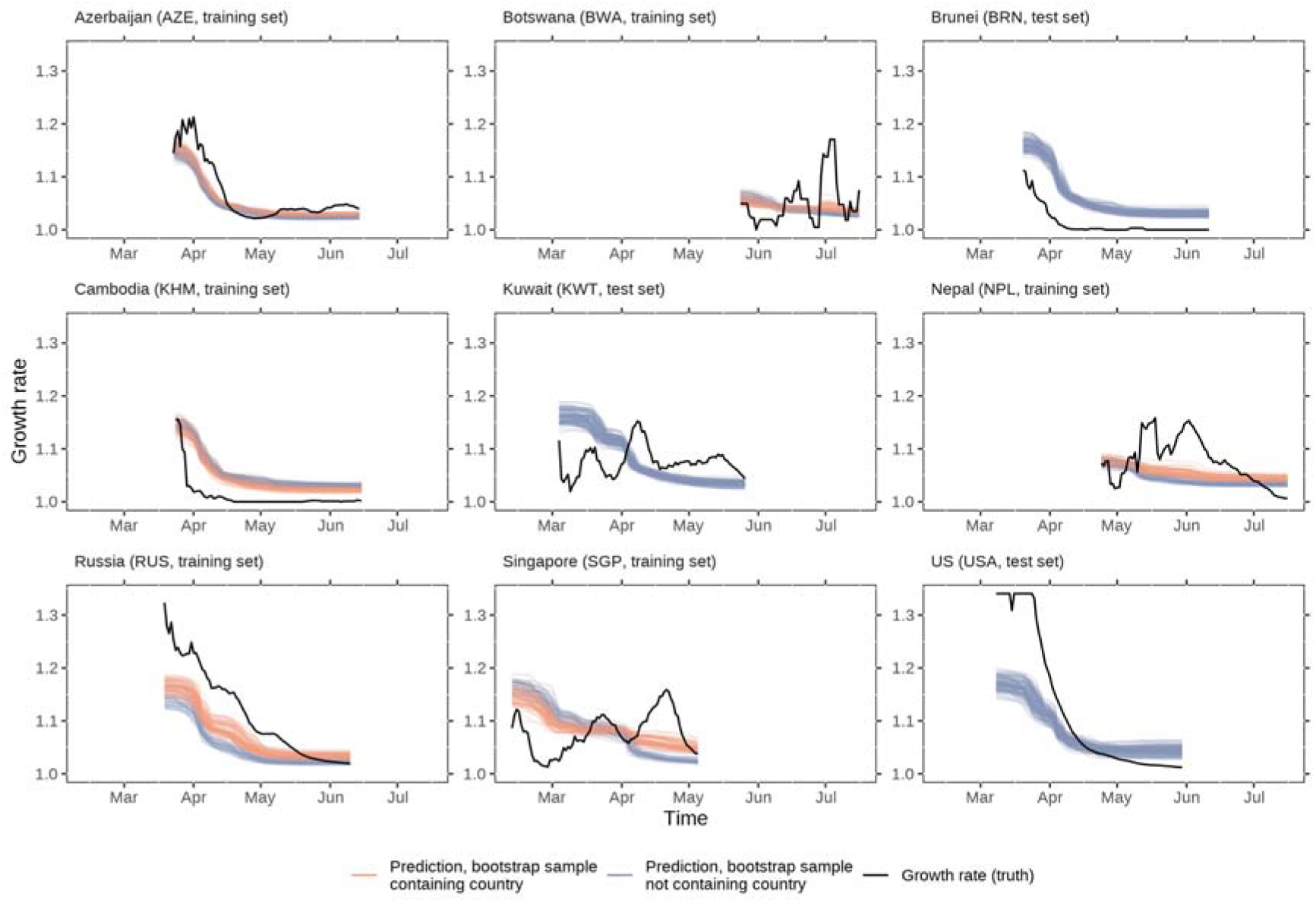
Country-specific growth rates and inaccurate predictions. The figure shows some examples of countries that the models’ predictions were inaccurate. For some countries, the predicted growth rate was systematically higher (e.g., BRN, KHM) or lower (e.g., RUS) than the actual growth rate. In some instances, the reduction in the growth rate was predicted too early (e.g., AZE) or too late (e.g., KHM, BRN). The models were not able to predict very high growth rates in the beginning of the outbreak (e.g., USA, RUS), nor were they able to capture time series with untypical trends in the growth rate (e.g., BWA, KWT, NPL, SGP). Countries that were part of the training set contain bootstrap estimates of the predictions where the respective country was part of the bootstrap sample or not (indicated by the colour of the bootstrap estimates), countries that were in the test set contain only bootstrap estimates that did not contain the respective country.

## Supplementary Information

**Supplementary Tables**

**Supplementary Table 1.**
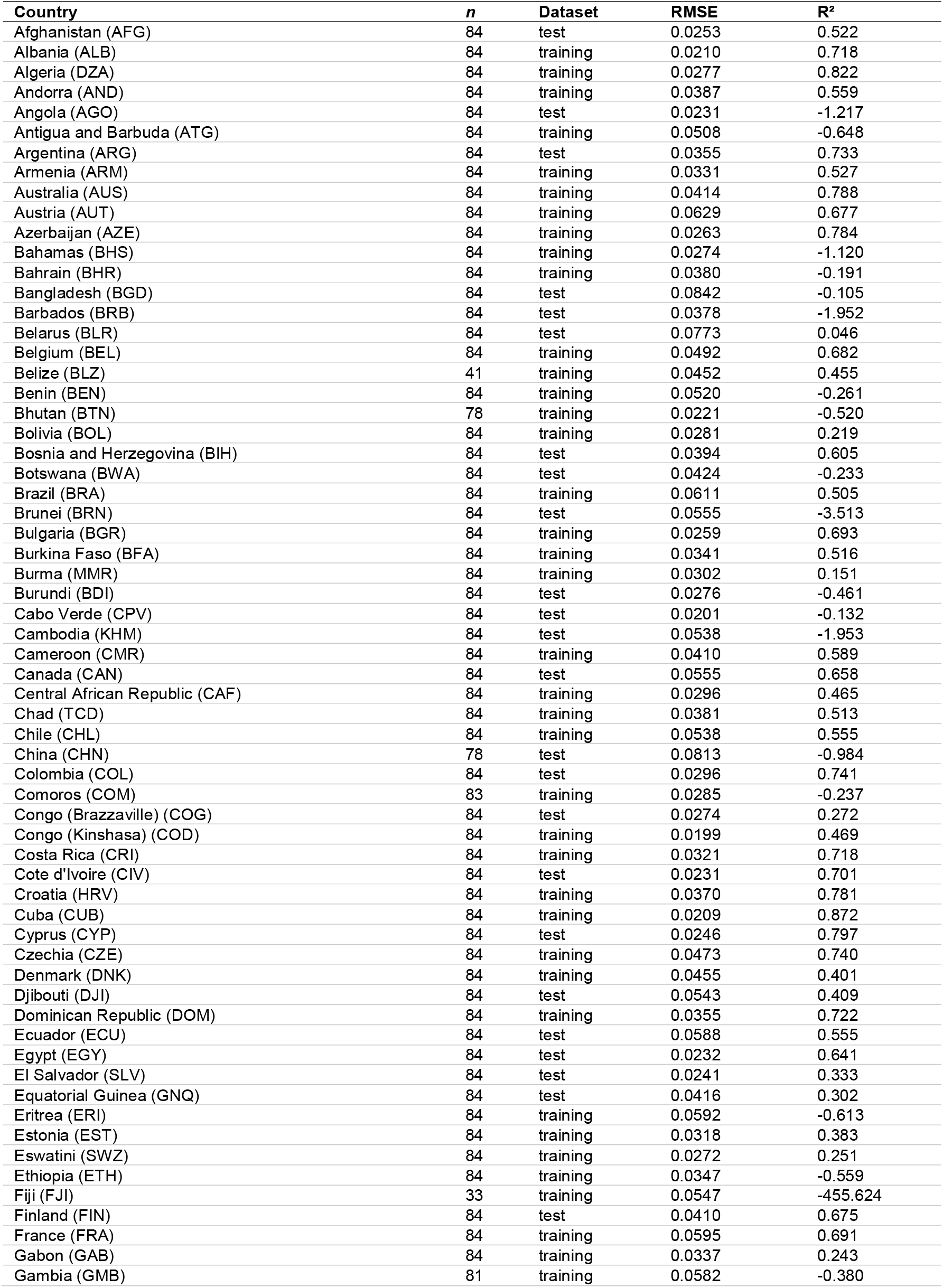
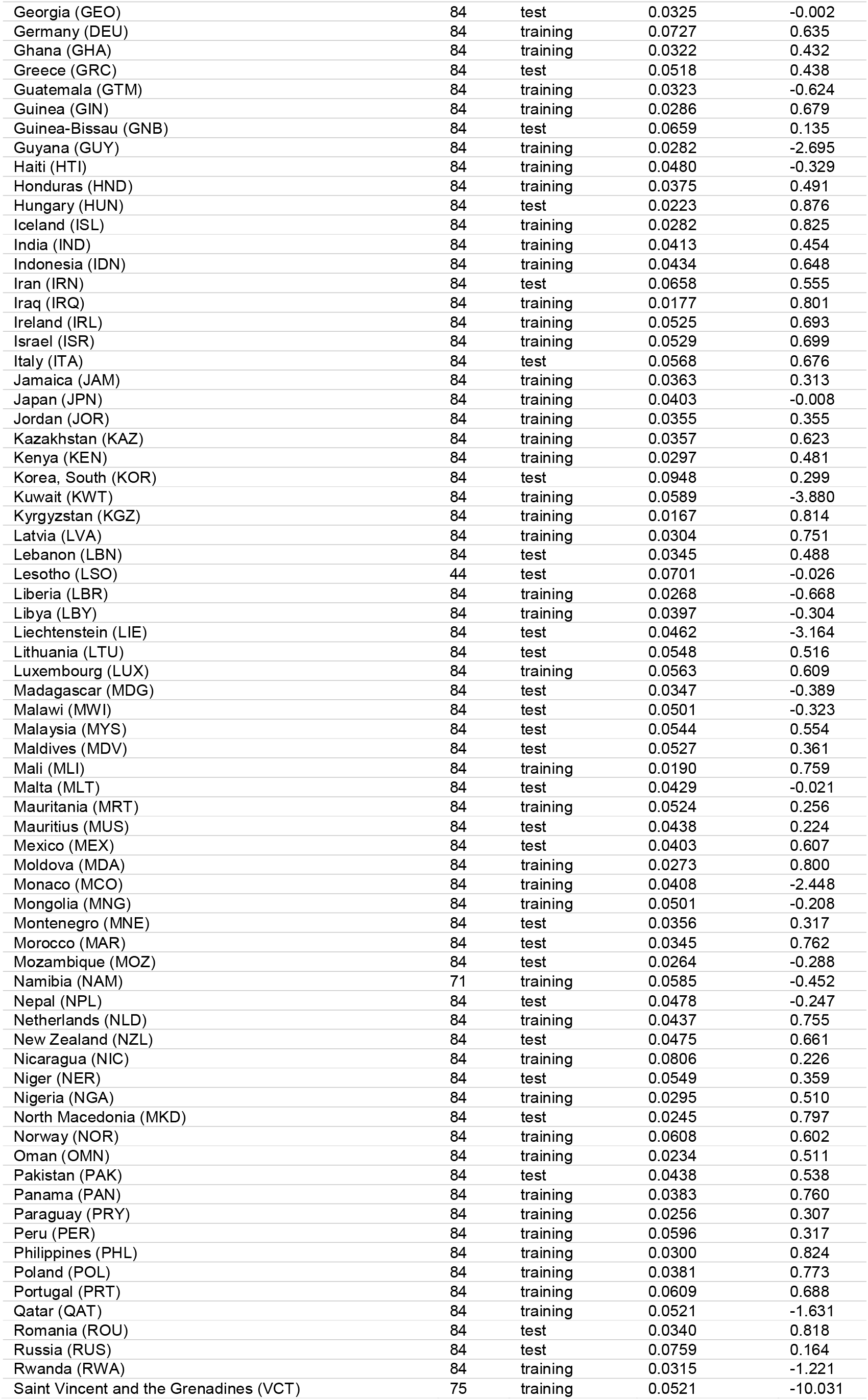
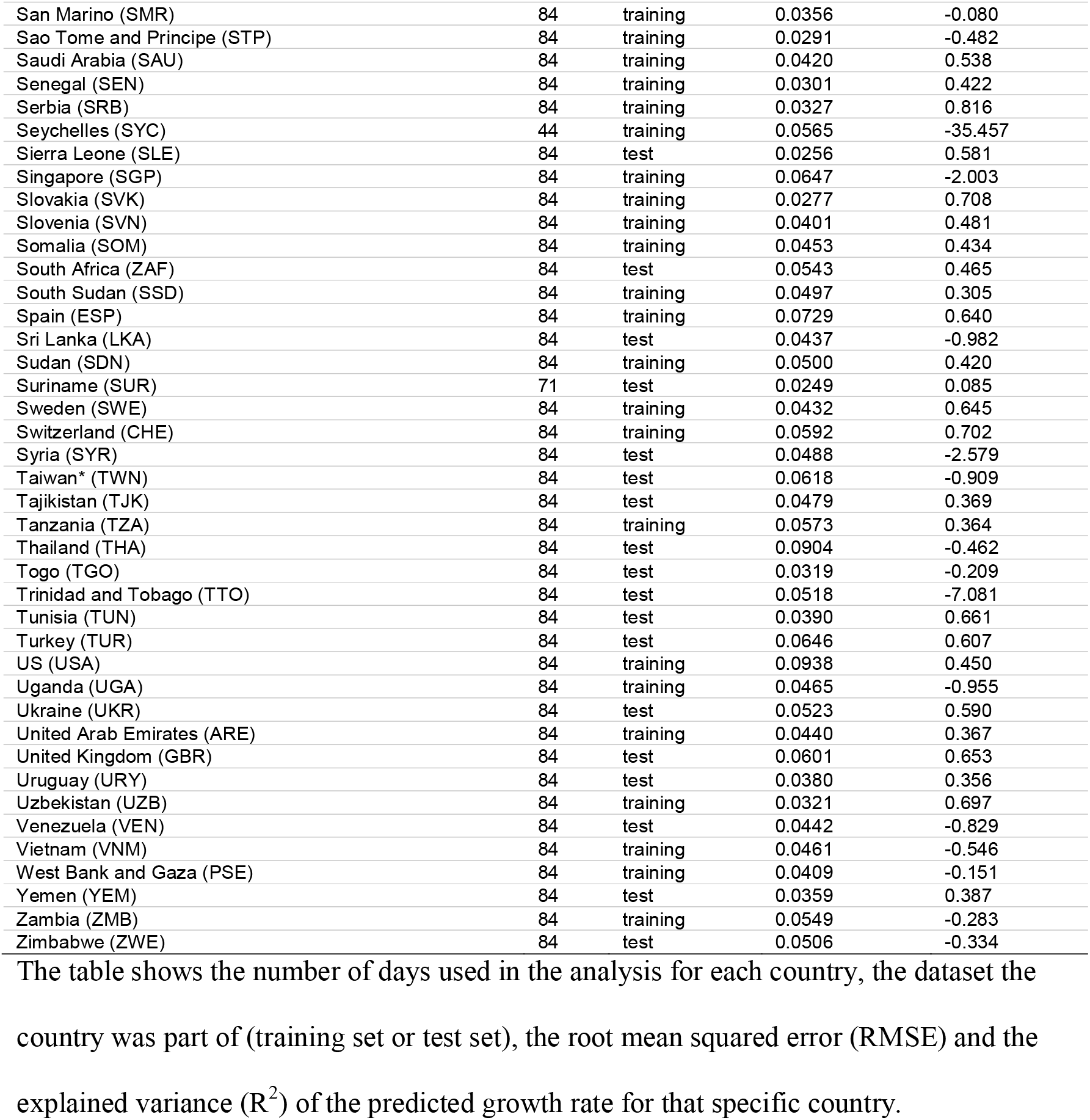
Countries included in the analysis

| Country | *n* | Dataset | RMSE | R<sup>2</sup> |
| --- | --- | --- | --- | --- |
| Afghanistan (AFG) | 84 | test | 0.0253 | 0.522 |
| Albania (ALB) | 84 | training | 0.0210 | 0.718 |
| Algeria (DZA) | 84 | training | 0.0277 | 0.822 |
| Andorra (AND) | 84 | training | 0.0387 | 0.559 |
| Angola (AGO) | 84 | test | 0.0231 | -1.217 |
| Antigua and Barbuda (ATG) | 84 | training | 0.0508 | -0.648 |
| Argentina (ARG) | 84 | test | 0.0355 | 0.733 |
| Armenia (ARM) | 84 | training | 0.0331 | 0.527 |
| Australia (AUS) | 84 | training | 0.0414 | 0.788 |
| Austria (AUT) | 84 | training | 0.0629 | 0.677 |
| Azerbaijan (AZE) | 84 | training | 0.0263 | 0.784 |
| Bahamas (BHS) | 84 | training | 0.0274 | -1.120 |
| Bahrain (BHR) | 84 | training | 0.0380 | -0.191 |
| Bangladesh (BGD) | 84 | test | 0.0842 | -0.105 |
| Barbados (BRB) | 84 | test | 0.0378 | -1.952 |
| Belarus (BLR) | 84 | test | 0.0773 | 0.046 |
| Belgium (BEL) | 84 | training | 0.0492 | 0.682 |
| Belize (BLZ) | 41 | training | 0.0452 | 0.455 |
| Benin (BEN) | 84 | training | 0.0520 | -0.261 |
| Bhutan (BTN) | 78 | training | 0.0221 | -0.520 |
| Bolivia (BOL) | 84 | training | 0.0281 | 0.219 |
| Bosnia and Herzegovina (BIH) | 84 | test | 0.0394 | 0.605 |
| Botswana (BWA) | 84 | test | 0.0424 | -0.233 |
| Brazil (BRA) | 84 | training | 0.0611 | 0.505 |
| Brunei (BRN) | 84 | test | 0.0555 | -3.513 |
| Bulgaria (BGR) | 84 | training | 0.0259 | 0.693 |
| Burkina Faso (BFA) | 84 | training | 0.0341 | 0.516 |
| Burma (MMR) | 84 | training | 0.0302 | 0.151 |
| Burundi (BDI) | 84 | test | 0.0276 | -0.461 |
| Cabo Verde (CPV) | 84 | test | 0.0201 | -0.132 |
| Cambodia (KHM) | 84 | test | 0.0538 | -1.953 |
| Cameroon (CMR) | 84 | training | 0.0410 | 0.589 |
| Canada (CAN) | 84 | test | 0.0555 | 0.658 |
| Central African Republic (CAF) | 84 | training | 0.0296 | 0.465 |
| Chad (TCD) | 84 | training | 0.0381 | 0.513 |
| Chile (CHL) | 84 | training | 0.0538 | 0.555 |
| China (CHN) | 78 | test | 0.0813 | -0.984 |
| Colombia (COL) | 84 | test | 0.0296 | 0.741 |
| Comoros (COM) | 83 | training | 0.0285 | -0.237 |
| Congo (Brazzaville) (COG) | 84 | test | 0.0274 | 0.272 |
| Congo (Kinshasa) (COD) | 84 | training | 0.0199 | 0.469 |
| Costa Rica (CRI) | 84 | training | 0.0321 | 0.718 |
| Cote d'Ivoire (CIV) | 84 | test | 0.0231 | 0.701 |
| Croatia (HRV) | 84 | training | 0.0370 | 0.781 |
| Cuba (CUB) | 84 | training | 0.0209 | 0.872 |
| Cyprus (CYP) | 84 | test | 0.0246 | 0.797 |
| Czechia (CZE) | 84 | training | 0.0473 | 0.740 |
| Denmark (DNK) | 84 | training | 0.0455 | 0.401 |
| Djibouti (DJI) | 84 | test | 0.0543 | 0.409 |
| Dominican Republic (DOM) | 84 | training | 0.0355 | 0.722 |
| Ecuador (ECU) | 84 | test | 0.0588 | 0.555 |
| Egypt (EGY) | 84 | test | 0.0232 | 0.641 |
| El Salvador (SLV) | 84 | test | 0.0241 | 0.333 |
| Equatorial Guinea (GNQ) | 84 | test | 0.0416 | 0.302 |
| Eritrea (ERI) | 84 | training | 0.0592 | -0.613 |
| Estonia (EST) | 84 | training | 0.0318 | 0.383 |
| Eswatini (SWZ) | 84 | training | 0.0272 | 0.251 |
| Ethiopia (ETH) | 84 | training | 0.0347 | -0.559 |
| Fiji (FJI) | 33 | training | 0.0547 | -455.624 |
| Finland (FIN) | 84 | test | 0.0410 | 0.675 |
| France (FRA) | 84 | training | 0.0595 | 0.691 |
| Gabon (GAB) | 84 | training | 0.0337 | 0.243 |
| Gambia (GMB) | 81 | training | 0.0582 | -0.380 |
| Georgia (GEO) | 84 | test | 0.0325 | -0.002 |
| Germany (DEU) | 84 | training | 0.0727 | 0.635 |
| Ghana (GHA) | 84 | training | 0.0322 | 0.432 |
| Greece (GRC) | 84 | test | 0.0518 | 0.438 |
| Guatemala (GTM) | 84 | training | 0.0323 | -0.624 |
| Guinea (GIN) | 84 | training | 0.0286 | 0.679 |
| Guinea-Bissau (GNB) | 84 | test | 0.0659 | 0.135 |
| Guyana (GUY) | 84 | training | 0.0282 | -2.695 |
| Haiti (HTI) | 84 | training | 0.0480 | -0.329 |
| Honduras (HND) | 84 | training | 0.0375 | 0.491 |
| Hungary (HUN) | 84 | test | 0.0223 | 0.876 |
| Iceland (ISL) | 84 | training | 0.0282 | 0.825 |
| India (IND) | 84 | training | 0.0413 | 0.454 |
| Indonesia (IDN) | 84 | training | 0.0434 | 0.648 |
| Iran (IRN) | 84 | test | 0.0658 | 0.555 |
| Iraq (IRQ) | 84 | training | 0.0177 | 0.801 |
| Ireland (IRL) | 84 | training | 0.0525 | 0.693 |
| Israel (ISR) | 84 | training | 0.0529 | 0.699 |
| Italy (ITA) | 84 | test | 0.0568 | 0.676 |
| Jamaica (JAM) | 84 | training | 0.0363 | 0.313 |
| Japan (JPN) | 84 | training | 0.0403 | -0.008 |
| Jordan (JOR) | 84 | training | 0.0355 | 0.355 |
| Kazakhstan (KAZ) | 84 | training | 0.0357 | 0.623 |
| Kenya (KEN) | 84 | training | 0.0297 | 0.481 |
| Korea, South (KOR) | 84 | test | 0.0948 | 0.299 |
| Kuwait (KWT) | 84 | training | 0.0589 | -3.880 |
| Kyrgyzstan (KGZ) | 84 | training | 0.0167 | 0.814 |
| Latvia (LVA) | 84 | training | 0.0304 | 0.751 |
| Lebanon (LBN) | 84 | test | 0.0345 | 0.488 |
| Lesotho (LSO) | 44 | test | 0.0701 | -0.026 |
| Liberia (LBR) | 84 | training | 0.0268 | -0.668 |
| Libya (LBY) | 84 | training | 0.0397 | -0.304 |
| Liechtenstein (LIE) | 84 | test | 0.0462 | -3.164 |
| Lithuania (LTU) | 84 | test | 0.0548 | 0.516 |
| Luxembourg (LUX) | 84 | training | 0.0563 | 0.609 |
| Madagascar (MDG) | 84 | test | 0.0347 | -0.389 |
| Malawi (MWI) | 84 | test | 0.0501 | -0.323 |
| Malaysia (MYS) | 84 | test | 0.0544 | 0.554 |
| Maldives (MDV) | 84 | test | 0.0527 | 0.361 |
| Mali (MLI) | 84 | training | 0.0190 | 0.759 |
| Malta (MLT) | 84 | test | 0.0429 | -0.021 |
| Mauritania (MRT) | 84 | training | 0.0524 | 0.256 |
| Mauritius (MUS) | 84 | test | 0.0438 | 0.224 |
| Mexico (MEX) | 84 | test | 0.0403 | 0.607 |
| Moldova (MDA) | 84 | training | 0.0273 | 0.800 |
| Monaco (MCO) | 84 | training | 0.0408 | -2.448 |
| Mongolia (MNG) | 84 | training | 0.0501 | -0.208 |
| Montenegro (MNE) | 84 | test | 0.0356 | 0.317 |
| Morocco (MAR) | 84 | test | 0.0345 | 0.762 |
| Mozambique (MOZ) | 84 | test | 0.0264 | -0.288 |
| Namibia (NAM) | 71 | training | 0.0585 | -0.452 |
| Nepal (NPL) | 84 | test | 0.0478 | -0.247 |
| Netherlands (NLD) | 84 | training | 0.0437 | 0.755 |
| New Zealand (NZL) | 84 | test | 0.0475 | 0.661 |
| Nicaragua (NIC) | 84 | training | 0.0806 | 0.226 |
| Niger (NER) | 84 | test | 0.0549 | 0.359 |
| Nigeria (NGA) | 84 | training | 0.0295 | 0.510 |
| North Macedonia (MKD) | 84 | test | 0.0245 | 0.797 |
| Norway (NOR) | 84 | training | 0.0608 | 0.602 |
| Oman (OMN) | 84 | training | 0.0234 | 0.511 |
| Pakistan (PAK) | 84 | test | 0.0438 | 0.538 |
| Panama (PAN) | 84 | training | 0.0383 | 0.760 |
| Paraguay (PRY) | 84 | training | 0.0256 | 0.307 |
| Peru (PER) | 84 | training | 0.0596 | 0.317 |
| Philippines (PHL) | 84 | training | 0.0300 | 0.824 |
| Poland (POL) | 84 | training | 0.0381 | 0.773 |
| Portugal (PRT) | 84 | training | 0.0609 | 0.688 |
| Qatar (QAT) | 84 | training | 0.0521 | -1.631 |
| Romania (ROU) | 84 | test | 0.0340 | 0.818 |
| Russia (RUS) | 84 | test | 0.0759 | 0.164 |
| Rwanda (RWA) | 84 | training | 0.0315 | -1.221 |
| Saint Vincent and the Grenadines (VCT) | 75 | training | 0.0521 | -10.031 |
| San Marino (SMR) | 84 | training | 0.0356 | -0.080 |
| Sao Tome and Principe (STP) | 84 | training | 0.0291 | -0.482 |
| Saudi Arabia (SAU) | 84 | training | 0.0420 | 0.538 |
| Senegal (SEN) | 84 | training | 0.0301 | 0.422 |
| Serbia (SRB) | 84 | training | 0.0327 | 0.816 |
| Seychelles (SYC) | 44 | training | 0.0565 | -35.457 |
| Sierra Leone (SLE) | 84 | test | 0.0256 | 0.581 |
| Singapore (SGP) | 84 | training | 0.0647 | -2.003 |
| Slovakia (SVK) | 84 | training | 0.0277 | 0.708 |
| Slovenia (SVN) | 84 | training | 0.0401 | 0.481 |
| Somalia (SOM) | 84 | training | 0.0453 | 0.434 |
| South Africa (ZAF) | 84 | test | 0.0543 | 0.465 |
| South Sudan (SSD) | 84 | training | 0.0497 | 0.305 |
| Spain (ESP) | 84 | training | 0.0729 | 0.640 |
| Sri Lanka (LKA) | 84 | test | 0.0437 | -0.982 |
| Sudan (SDN) | 84 | training | 0.0500 | 0.420 |
| Suriname (SUR) | 71 | test | 0.0249 | 0.085 |
| Sweden (SWE) | 84 | training | 0.0432 | 0.645 |
| Switzerland (CHE) | 84 | training | 0.0592 | 0.702 |
| Syria (SYR) | 84 | test | 0.0488 | -2.579 |
| Taiwan* (TWN) | 84 | test | 0.0618 | -0.909 |
| Tajikistan (TJK) | 84 | test | 0.0479 | 0.369 |
| Tanzania (TZA) | 84 | training | 0.0573 | 0.364 |
| Thailand (THA) | 84 | test | 0.0904 | -0.462 |
| Togo (TGO) | 84 | test | 0.0319 | -0.209 |
| Trinidad and Tobago (TTO) | 84 | test | 0.0518 | -7.081 |
| Tunisia (TUN) | 84 | test | 0.0390 | 0.661 |
| Turkey (TUR) | 84 | test | 0.0646 | 0.607 |
| US (USA) | 84 | training | 0.0938 | 0.450 |
| Uganda (UGA) | 84 | training | 0.0465 | -0.955 |
| Ukraine (UKR) | 84 | test | 0.0523 | 0.590 |
| United Arab Emirates (ARE) | 84 | training | 0.0440 | 0.367 |
| United Kingdom (GBR) | 84 | test | 0.0601 | 0.653 |
| Uruguay (URY) | 84 | test | 0.0380 | 0.356 |
| Uzbekistan (UZB) | 84 | training | 0.0321 | 0.697 |
| Venezuela (VEN) | 84 | test | 0.0442 | -0.829 |
| Vietnam (VNM) | 84 | training | 0.0461 | -0.546 |
| West Bank and Gaza (PSE) | 84 | training | 0.0409 | -0.151 |
| Yemen (YEM) | 84 | test | 0.0359 | 0.387 |
| Zambia (ZMB) | 84 | training | 0.0549 | -0.283 |
| Zimbabwe (ZWE) | 84 | test | 0.0506 | -0.334 |

The table shows the number of days used in the analysis for each country, the dataset the country was part of (training set or test set), the root mean squared error (RMSE) and the explained variance ( $R^2$ ) of the predicted growth rate for that specific country.

## Supplementary Notes

### Learning from the data

In contrast to analysing the cumulative confirmed COVID-19 cases for individual countries as individual time series, the approach used in this study aims at estimating the mean effect of a non-pharmaceutical intervention (NPI) over all countries that have implemented it. The effect is quantified by how much the current growth rate at a specific point in time (relative to the implementation date) differs between countries that have implemented the measure vs. countries that have not implemented it, as well as how it differs at specific times after implementation (i.e., the growth rate is expected to be influenced differently by an NPI on the first day after implementation compared to the second or third day, etc.). Assuming the growth rate to be constant during the initial phase of exponential growth with no NPIs in place, a lower-than-normal growth rate should co-occur with an NPI being in place, or, more realistically, a set of NPIs being in place at the same time. This assumption is expected to be valid both for the time domain, as well as geographically. It should not matter in which country these measures are in place, as we are only interested in an average effect of these NPIs. This allows the machine learning model to disentangle the effects of different NPIs, as different countries have implemented different subsets of all possible NPIs, However, NPIs that have been implemented later, after other NPIs already have been in place, might start from a lower growth rate, hence will have less chance to reduce it. This concern is more severe if different countries have implemented a similar set of NPIs in similar sequence, and less severe if they have done so in different sequence with different timing during the spread of the COVID-19 pandemic, which will be investigated in the following paragraphs.

Within each country, the features representing the NPIs are highly correlated (all Pearson correlations *r* > .69, with a median correlation of *r* = .96), because a country has either not implemented a pair of measures (resulting in a correlation of one), or it has implemented two measures in close succession, leading to a very high correlation. However, in the model we do not distinguish between different countries. We want to derive the average effect over all countries. We assume that the dependent variable in the model, the growth rate, covaries with an NPI being in place irrespective of which country that data point has originated from. Hence, as long as different countries have implemented sufficiently different subsets of NPIs, the model can estimate the effect of an NPI. Different countries having implemented different subsets of NPIs would result in lower correlations of the NPI-related features across all countries.

Another characteristic that can help the model learn the effects of different NPIs is the chronological order that different countries have implemented different NPIs. Different chronological order in different countries would also result in lower correlations of the measures across all countries. And the correlations are indeed lower when calculated across (and not within) countries (minimum Pearson correlation *r* = −0.42, median correlation of *r* = 0.14, maximum at *r* = 0.72), substantiating the assumption that different countries have implemented different subsets and/or have implemented the NPIs in different chronological order.

### Additional discussion of results: Other NPIs

Other NPIs used in the analysis were associated with only minor effects (see Extended Data Fig. 2-5 for plots of all NPIs). This includes lockdown and curfews, which were, on average, used rather late in the outbreak within each country (Extended Data Fig. 1). Despite being associated with minor average effects, some NPIs showed high variation in the bootstrap estimates of their effects. Examples are inbound external border restrictions, voluntary social distancing, and restriction and regulation of government services. Hence, globally, the effects were not significant, but this does not rule out effects in specific countries or under specific circumstances.

There could be multiple reasons for these weak effects. Most importantly, the strategies that are reported in the CoronaNet dataset are not mutually exclusive. For example, restricting mass gatherings reduces the need for travel, which in turn reduces the potential impact of internal and external border restrictions. Additionally, NPIs that have been implemented later, after other NPIs already have been in place, might start from a lower growth rate, hence will have less chance to reduce it. This is especially true if different countries have implemented a similar set of NPIs in similar sequence. The data shows that there is variation in the timing and sequence of NPI implementation across countries (see supplementary material), which at least reduces this concern to some extent (see above).

Also, NPIs that are in effect for a very short timeframe, like curfews or lockdowns that last for only a few days, might have only minor effects which are hard to detect. Similar concerns apply for NPIs on subnational level, which affect fewer people by definition.

### Additional discussion of results: Country-specific covariates

The relations of country-specific covariates and predicted growth rate were relatively small compared to NPIs and time-related, NPI-independent effects, and the random variation in the bootstrap samples was large (Fig. 3). The largest effect in this group of covariates was the percentage of the population aged 65 and above. Countries with a lower percentage, i.e., countries with a younger population, showed slightly higher growth rates. The effect was marginally larger than the bootstrap variation.

Countries with high low GDP (ppp) per capita seemed to have slightly higher growth rates on average, but this effect did not exceed the bootstrap variation. For the percentage of people being exposed to high levels of air pollution and the percentage of urban population, there was no significant median effect. However, the bootstraps samples for the latter covariate show very high variation in the extremes (very high or very low percentage), compared to the median ranges. This might indicate differences in the average growth rate within the countries with very high or low percentage of urban population.

## References and Notes

1 WHO. Coronavirus disease 2019 (COVID-19) situation report 97. 2020. https://www.who.int/docs/default-source/coronaviruse/situation-reports/20200426-sitrep97-covid-19.pdf?sfvrsn=d1c3e800_6 (accessed April 26, 2020).

2 Bedford J, Enria D, Giesecke J, et al. COVID-19: towards controlling of a pandemic. The Lancet 2020; 395: 1015–8.

3 Nussbaumer-Streit B, Mayr V, Dobrescu AI, et al. Quarantine alone or in combination with other public health measures to control COVID19: a rapid review. Cochrane Database Syst Rev 2020. DOI:10.1002/14651858.CD013574.

4 Viner RM, Russell SJ, Croker H, et al. School closure and management practices during coronavirus outbreaks including COVID-19: a rapid systematic review. Lancet Child Adolesc Health 2020; 4: 397–404.

5 Maier BF, Brockmann D. Effective containment explains subexponential growth in recent confirmed COVID-19 cases in China. Science 2020; 368: 742–6.

6 Prem K, Liu Y, Russell TW, et al. The effect of control strategies to reduce social mixing on outcomes of the COVID-19 epidemic in Wuhan, China: a modelling study. Lancet Public Health 2020; 5: e261–70.

7 Lai S, Ruktanonchai NW, Zhou L, et al. Effect of non-pharmaceutical interventions to contain COVID-19 in China. Nature 2020;: 1–5.

8 Pan A, Liu L, Wang C, et al. Association of public health interventions with the epidemiology of the COVID-19 outbreak in Wuhan, China. JAMA 2020; 323: 1915–23.

9 Hsiang S, Allen D, Annan-Phan S, et al. The effect of large-scale anti-contagion policies on the COVID-19 pandemic. Nature 2020;: 1–9.

10 Flaxman S, Mishra S, Gandy A, et al. Estimating the effects of non-pharmaceutical interventions on COVID-19 in Europe. Nature 2020;: 1–5.

11 Brauner JM, Mindermann S, Sharma M, et al. The effectiveness of eight nonpharmaceutical interventions against COVID-19 in 41 countries. *medRxiv* 2020;: 2020.05.28.20116129.

12 Novel Coronavirus (COVID-19) Cases Data - Humanitarian Data Exchange. https://data.humdata.org/dataset/novel-coronavirus-2019-ncov-cases (accessed April 23, 2020).

13 Cheng C, Barceló J, Hartnett AS, Kubinec R, Messerschmidt L. COVID-19 Government Response Event Dataset (CoronaNet v.1.0). Nat Hum Behav 2020; 4: 756–68.

14 Piburn J. wbstats: Programmatic Access to the World Bank API. Oak Ridge, Tennessee: Oak Ridge National Laboratory, 2018 https://www.ornl.gov/division/csed/gist.

15 Wilder B, Charpignon M, Killian JA, et al. Modeling Between-Population Variation in COVID-19 Dynamics in Hubei, Lombardy, and New York City. Rochester, NY: Social Science Research Network, 2020 DOI:10.2139/ssrn.3564800.

16 Population ages 65 and above (% of total population) | Data. https://data.worldbank.org/indicator/SP.POP.65UP.TO.ZS (accessed July 28, 2020).

17 Rao ASRS, Vazquez JA. Identification of COVID-19 can be quicker through artificial intelligence framework using a mobile phone-based survey in the populations when cities/towns are under quarantine. Infect Control Hosp Epidemiol 2020. DOI:10.1017/ice.2020.61.

18 Population, total | Data. https://data.worldbank.org/indicator/SP.POP.TOTL (accessed July 28, 2020).

19 Urban population | Data. https://data.worldbank.org/indicator/SP.URB.TOTL (accessed July 28, 2020).

20 Peng L, Zhao X, Tao Y, Mi S, Huang J, Zhang Q. The effects of air pollution and meteorological factors on measles cases in Lanzhou, China. Environ Sci Pollut Res 2020; 27: 13524–33.

21 Fattorini D, Regoli F. Role of the chronic air pollution levels in the Covid-19 outbreak risk in Italy. Environ Pollut 2020; 264: 114732.

22 PM2.5 air pollution, population exposed to levels exceeding WHO guideline value (% of total) | Data. https://data.worldbank.org/indicator/EN.ATM.PM25.MC.ZS (accessed July 28, 2020).

23 Wright AL, Sonin K, Driscoll J, Wilson J. Poverty and Economic Dislocation Reduce Compliance with COVID-19 Shelter-in-Place Protocols. Rochester, NY: Social Science Research Network, 2020 DOI:10.2139/ssrn.3573637.

24 GDP per capita, PPP (current international $) | Data. https://data.worldbank.org/indicator/NY.GDP.PCAP.PP.CD?end=2019&start=1990 (accessed July 28, 2020).

25 Jung S, Akhmetzhanov AR, Hayashi K, et al. Real-time estimation of the risk of death from novel coronavirus (COVID-19) infection: inference using exported cases. J Clin Med 2020; 9: 523.

26 Ma J. Estimating epidemic exponential growth rate and basic reproduction number. Infect Dis Model 2020; 5: 129–41.

27 Muniz-Rodriguez K, Chowell G, Cheung C-H, et al. Doubling Time of the COVID-19 Epidemic by Chinese Province. *medRxiv* 2020; published online April 24. DOI:10.1101/2020.02.05.20020750.

28 WHO. Coronavirus disease 2019 (COVID-19) situation report 12 (Indonesia). 2020. https://www.who.int/docs/default-source/searo/indonesia/covid19/who-situation-report12.pdf?sfvrsn=811c7f19_2 (accessed July 28, 2020).

29 WHO. Coronavirus disease 2019 (COVID-19) situation report 183. 2020. https://www.who.int/docs/default-source/wha-70-and-phe/20200721-covid-19-sitrep183.pdf?sfvrsn=b3869b3_2 (accessed July 28, 2020).

30 Singer HM. The COVID-19 pandemic: growth patterns, power law scaling, and saturation. Phys Biol 2020. DOI:10.1088/1478-3975/ab9bf5.

31 Brouwer ED, Raimondi D, Moreau Y. Modeling the COVID-19 outbreaks and the effectiveness of the containment measures adopted across countries. *medRxiv* 2020; published online April 10. DOI:10.1101/2020.04.02.20046375.

32 Harper CA, Satchell LP, Fido D, Latzman RD. Functional Fear Predicts Public Health Compliance in the COVID-19 Pandemic. Int J Ment Health Addict 2020;: 1–14.

33 Dryhurst S, Schneider CR, Kerr J, et al. Risk perceptions of COVID-19 around the world. J Risk Res 2020; 0: 1–13.

34 Wright MN, Ziegler A. ranger: A Fast Implementation of Random Forests for High Dimensional Data in C++ and R. J Stat Softw 2017; 77. DOI:10.18637/jss.v077.i01.

35 Bergstra J, Bengio Y. Random Search for Hyper-Parameter Optimization. J Mach Learn Res 2012; 13: 281–305.

36 Apley DW, Zhu J. Visualizing the effects of predictor variables in black box supervised learning models. ArXiv161208468 Stat 2019; published online Aug 19. http://arxiv.org/abs/1612.08468 (accessed April 20, 2020).

37 Molnar C. Interpretable Machine Learning. https://christophm.github.io/interpretable-mlbook/ (accessed April 20, 2020).

38 R Core Team. R: A Language and Environment for Statistical Computing. Vienna, Austria: R Foundation for Statistical Computing, 2020 https://www.R-project.org/.

39 Lang M, Binder M, Richter J, et al. mlr3: A modern object-oriented machine learning framework in R. J Open Source Softw 2019; 4: 1903.

40 Molnar C. iml: An R package for Interpretable Machine Learning. J Open Source Softw 2018; 3: 786.

41 Wickham H. ggplot2: Elegant Graphics for Data Analysis. Springer-Verlag New York, 2016 https://ggplot2.tidyverse.org.

42 Ferguson N, Laydon D, Nedjati Gilani G, et al. Report 9: Impact of non-pharmaceutical interventions (NPIs) to reduce COVID19 mortality and healthcare demand. 2020 DOI:10.25561/77482.

43 Auger KA, Shah SS, Richardson T, et al. Association Between Statewide School Closure and COVID-19 Incidence and Mortality in the US. JAMA 2020; published online July 29. DOI:10.1001/jama.2020.14348.

44 Williams SN, Armitage CJ, Tampe T, Dienes K. Public perceptions and experiences of social distancing and social isolation during the COVID-19 pandemic: a UK-based focus group study. BMJ Open 2020; 10: e039334.

45 Yuan J, Li M, Lv G, Lu ZK. Monitoring transmissibility and mortality of COVID-19 in Europe. Int J Infect Dis 2020; 95: 311–5.

46 Tang JW. The effect of environmental parameters on the survival of airborne infectious agents. J R Soc Interface 2009; 6: S737–46.

47 Tsang TK, Wu P, Lin Y, Lau EHY, Leung GM, Cowling BJ. Effect of changing case definitions for COVID-19 on the epidemic curve and transmission parameters in mainland China: a modelling study. Lancet Public Health 2020; 0. DOI:10.1016/S24682667(20)30089-X.

